# Immune profile and responses of a novel Dengue DNA vaccine encoding EDIII-NS1 consensus design based on Indo-African sequences

**DOI:** 10.1101/2021.09.21.21263883

**Authors:** Arun Sankaradoss, Suraj Jagtap, Junaid Nazir, Shefta E-Moula, Ayan Modak, Joshuah Fialho, Meenakshi Iyer, Jayanthi S Shastri, Mary Dias, Ravisekhar Gadepalli, Alisha Aggarwal, Manoj Vedpathak, Sachee Agrawal, Awadhesh Pandit, Amul Nisheetha, Anuj Kumar, Mohamed Shafi, Swathi Balachandra, Tina Damodar, Moses Muia Masika, Patrick Mwaura, Omu Anzala, Kar Muthumani, Ramanathan Sowdhamini, Guruprasad R. Medigeshi, Rahul Roy, Chitra Pattabiraman, Sudhir Krishna, Easwaran Sreekumar

**Affiliations:** National Centre for Biological Sciences, Tata Institute of Fundamental Research, Bangalore, 560065, India; Department of Chemical Engineering, Indian Institute of Science, Bangalore, 560012, India; Molecular Virology Laboratory, Rajiv Gandhi Centre for Biotechnology (RGCB), Thiruvananthapuram, 695014, Kerala, India; Department of Microbiology, T.N.Medical College & B.y.L.Nair Hospital, Mumbai, 400008, India; Division of Infectious disease, St.John’s Medical College and Hospital, Bangalore, 560034, India; Department of Microbiology, All India Institute of Medical Sciences, Jodhpur, 342005, India; KAVI Institute of Clinical Research, University of Nairobi, Nairobi, 19676-00202, Kenya; Vaccine and Immunotherapy Center, Wistar Institute, Philadelphia, PA 19104, USA; Translational Health Science and Technology Institute, Faridabad, Haryana, 121001, India; Department of Chemical Engineering, Indian Institute of Science, Bangalore, India; Molecular Biophysics Unit, Indian Institute of Science, Bangalore, India; Center for Biosystems Science and Engineering, Indian Institute of Science, Bangalore, 560012, India; Department of Neurovirology, National Institute of Mental Health and Neurosciences, Bangalore, 560029, India; School of Interdisciplinary Life Sciences, Indian Institute of Technology Goa, Ponda, 404401, India

**Author notes:** Corresponding authors: Arun Sankaradoss,. Easwaran Sreekumar. Equal contribution (Suraj Jagtap, Junaid Nazir, Shefta E-Moula).

**Keywords:** Dengue, DNA vaccine, Antibody-dependent enhancement, Envelope, EDIII domain, NS1 protein

## Abstract

Following the recent clinical clearance of an Indian DNA COVID-19 vaccine, India and Africa are potential regions where DNA vaccines may become a major delivery system subject to a range of immunological and regulatory scrutiny. The ongoing COVID pandemic highlights the need to tackle viral variants and expand the number of antigens and assess diverse delivery systems. To address some of these key issues, we have created a Dengue DNA vaccine candidate with the EDIII region as the key antigen given the promise of this segment in not causing ADE, a challenge with this disease. In addition, we have added the NS1 region to broaden the immune response. Following a large Dengue viral sequencing exercise in India, complemented with data from east Africa, our approach was to generate a consensus of four serotypes ED3-NS1 vaccine to explore tackling the issue of diversity. Our *In silico* structural analysis of EDIII consensus vaccine sequence revealed that epitopes are structurally conserved and immunogenic across HLA diversity. Vaccination of mice with this construct induced pan-serotype neutralizing antibodies and antigen-specific T cell responses. Furthermore, the DNA vaccination confers protection against DENV challenge in AG129 mice. Finally, assaying of intracellular staining for IFN-γ, immunoglobulin IgG2(a/c) /IgG1 ratios as well as immune gene profiling suggested a strong Th1-dominant immune response. Our Dengue DNA platform with a focus on Indo-African sequences offers an approach for assessing cross reactive immunity in animal models and lays the foundation for human vaccine roll out either as a stand-alone or mix and match strategy.

## Introduction

Vaccine development is an evolving process. The current COVID-19 pandemic has facilitated the development of numerous vaccine technologies to mitigate public health crises. Among other technologies, nucleic acid vaccines have emerged as a rapid and versatile platform for an emergency, which is why these are among the very first COVID-19 vaccines in human use^1,2^. Nucleic acid vaccines require a short time from design to clinical trials, therefore, it may be possible to test together, in the same vaccine, different variants of antigens that cover circulating mutations^3^. Nucleic acid vaccines are adaptable for mix and match vaccination approaches. Recently, Barros-Martins et al. and Schmidt et al. reported that heterologous dose with SARS-CoV2 adenovirus-based vaccine (ChAdOx1) followed by SARS-CoV2 mRNA vaccine (BNT) induced a stronger immune response than homologous adenovirus vaccine regimen^4,5^. In addition, they offer more natural antigen presentation to the immune system, resulting in better T-cell responses^6^. An advantage of DNA over mRNA is they are more versatile, temperature stable, cost-effective, and cold chain free which are essential features for delivery to resource-limited settings^7–9^. After over 3 decades of research on DNA vaccines, the world’s first DNA vaccine against COVID-19 has been approved in India, for emergency use, highlighting the evolution of low-cost vaccine platforms.

In addition to the standard vaccine approaches (i.e. whole genome, peptide, and virus-like particle vaccines) several nucleic acid-based vaccines for DENV are in the pipeline^10–13^. DENV vaccine development is complicated by four antigenically diverse serotypes (DENV 1-4) and their intra-serotype (genotype) diversity at both local and global scales^14–16^. Several studies have reported the association between the genotype shift and magnitude of the outbreak and disease severity^17–22^. However, data on circulating genotype level information is limited in India. The authors’ data highlighted the emergence of DENV1 genotype I & V, DENV3 genotype III in Southern India and DENV2 cosmopolitan genotype dominated both in the South and in Delhi^23–25^. Previous studies in India reported co-circulation of DENV 4 genotype I strains in Pune, along with DENV 1, DENV2 and DENV 3^26,27^. Furthermore, Indian DENV sequences are close to African DENV’s particularly, Indian DENV2 cosmopolitan genotype found in various outbreaks in Kenya^28,29^.

The current DENV vaccine effort is also thought to be hampered by antibody-dependent enhancement (ADE), whereby cross-reactive antibodies against one serotype can enhance subsequent infection by heterologous serotype^30,31^. To circumvent such issues, regions or motifs of the antigen responsible for causing ADE must be eliminated from the vaccine design. Domain III of E protein has been identified as the major target of highly neutralizing and protective serotype-specific Abs while in contrast, PrM and EDI-II directed antibodies are reported to enhance infection *via* Antibody-dependent enhancement (ADE)^32–35^ . Recent studies have found that EDIII based DENV vaccines could circumvent ADE of infection in mice, whereas T cell response against EDIII DNA vaccine has shown to play a role in disease protection through effective viral clearance^36–38^. Several studies have also shown that EDIII directed antibodies can inhibit the entry of the flavivirus into target cells^39–44^. Mutations in EDIII may affect antibody binding as well as protein interaction with cellular receptors^23^. Previous studies have reported that mutations in EDIII of DENV and other flaviviruses have lower virulence or the ability to escape immune neutralization ^45^.

An effective DENV vaccine should be able to target circulating strain divergence; in this case, a consensus-based vaccine may be optimal. Consensus-based vaccines are an intriguing strategy which could minimize the sequence diversity across strains. Furthermore, recent studies on H1N1 and human immunodeficiency virus (HIV) in mice, have suggested that consensus immunogen-based vaccines may be ideal to address the viral genetic divergence^46,47^. Moreover, several studies have also shown that consensus-prediction could outperform single-sequence determination methods in vaccine design^48–50^.

DENV non-structural protein 1 (NS1) has also emerged as an attractive vaccine target. NS1, a highly conserved protein among flaviviruses, is involved in virus replication and immune evasion^51,52^. However, it has also been reported to activate antibody Fc-mediated effector function and provide protection against flaviviruses^53,54^ . NS1 has been demonstrated to trigger T-cell responses in both experimental animals and humans^51,52,55^ . Previous studies have shown that NS1 induced T cell responses involved in protection against dengue infection. Furthermore, vaccination with DENV1, DENV3, or DENV4 NS1 protected against a heterologous DENV2 challenge^54^. Taken together, combining EDIII and NS1 for vaccine design could be a possible way to circumvent immune interference among serotypes. Here, we further developed DENV EDIII based vaccines by integrating the sequence information from circulating genotypes of each serotype instead of a single strain genome sequence; incorporating NS1 to induce broad T cell responses; and then optimizing DNA vaccine construct for optimal protein expression.

This study employs whole-genome sequencing of DENV viruses from direct plasma/serum samples of dengue patients to profile contemporary DENV sequences across India. We examined DENV1-4 EDIII genetic variation in both local and global scales. Consequently, using the sequence data, Indian genotype-specific consensus-based polyvalent vaccine encoding DENV EDIII and NS1 was generated using a DNA vaccine platform. The immunogenicity of this vaccine candidate was evaluated by investigating its ability to induce tetravalent neutralizing antibodies, T cell responses as well as protective immunity in mice.

## Methods

### Samples and Ethical consideration

Serum/Plasma samples of DENV-NS1 positive patients were obtained from four hospitals in India viz. St. John’s medical college and hospital (SJRI), Bangalore, All India Institute of Medical Research (AIIMS), Jodhpur, Kasturba hospital for Infectious diseases (KHI), Mumbai, and All India Institute of Medical Research (AIIMS), Delhi during 2012-2018. The Institutional Ethical Clearance Review Boards approved the study of all institutions participating in this study (SJRI-IRB NO:-236/2016, AIIMS, Jodhpur-AIIMS/IEC/2017/49, & AIIMS/IEC/2019-20/939, KIH-IRB NO:07/2018, Ethics/THSTI/2011/2.1; AIIMS: IEC/NP:338/2011). Informed consent was obtained from enrolled patients. Clinical information of patients whose samples were included in this study was recorded in a preformed proforma which included the patient’s demographic details, signs, and symptoms, of illness, laboratory parameters, and treatment details.

### DENV viral RNA sequencing

Serum/ Plasma was tested for dengue virus-specific immunoglobulin M (IgM) and IgG antibodies using Panbio Dengue IgM and IgG capture ELISA kits, respectively (Catalog No. 01PE20/01PE21). Dengue infection classified as primary or secondary further samples were classified as per WHO 2009 guideline. According to the manufacturer’s instructions, Viral RNA was extracted from 150 µl of patient serum using QIAamp Viral RNA Mini Kit (Qiagen). Sequencing library preparation was performed with 1.2 ng of normalized ds cDNA using the Illumina Nextera XT sequencing kit and sequenced on an Illumina Miseq platform, using 2 × 75 and 2 × 150 bp paired-end reads. Raw sequence reads were inspected for quality using FASTQC (https://www.bioinformatics.babraham.ac.uk/projects/fastqc/), reads were filtered and trimmed based on the quality score.

Reads were then mapped to the reference sequences obtained from the RefSeq database (NC_001477(DENV1), NC_001474 (DENV2), NC_001475 (DENV3), NC_002640 (DENV4) using Geneious assembler (3 iterations and medium-low sensitivity). Assembled reads were checked manually for errors. Serotypes were assigned manually and coverage was assigned based on the following criteria: >10X coverage if the depth of the coverage at each base was >20 and breadth of the coverage >95%, 5X-10X coverage if depth <20 and breadth>95, 1X if depth<10 and breadth>90, for less coverages serotype. Consensus sequences were generated from the samples having >5X coverage based on the majority rule and were clipped to the length of reference sequences. iqtree v1.6.10 was used to construct maximum likelihood trees with 1000 bootstrap replicates. Genotypes were assigned to our sequences based on their positions in the tree. Figtree v1.4 was used to visualize the tree.

### DENV EDIII genetic diversity

Indian DENV sequences specific to the serotypes, including those sequenced during the study, were retrieved from the ViPR (Virus Pathogen Database and Analysis Resource) database. Sequence alignment was performed using MUSCLE and visualized in AliView. EDIII domain sequences were retained and used for downstream diversity analysis. EDIII sequences were compared pairwise against respective DENV reference strains and analyzed for Percentage diversity (Diversity%) and Percentage identity (Identity%). Diversity% was given by: (no. of variable sites/ EDIII length)*100 while Identity% was calculated as (100 - Diversity%).

### Dengue DNA vaccine construction

The polyvalent DENV vaccine DNA construct encodes ED III of all four serotypes and the NS1 sequence of DENV2. The consensus gene sequences were constructed using the predicted consensus sequences from sequences obtained from our study and Indian-specific sequences available in the ViPR. Multiple sequence alignment was performed using Geneious software. (https://mafft.cbrc.jp/alignment/server) and visualized in AliView.

The DENV DNA vaccine expression cassette was designed with the consensus sequences of the ED III gene of all four serotypes and consensus sequences of the NS1 gene of DENV2 linked together in a single construct with furin cleavage sites between the individual genes. Consensus sequences were optimized for ED III and NS1 expression, including codon and RNA optimization. The Kozak sequences and IgE leader sequence were added upstream to the DNA sequences as were furin cleavage sites to facilitate EDIII and NS1 processing. Finally, the synthetic DENV DNA expression cassette was inserted into the pVAX1 expression vector (Invitrogen, CA) between NheI and HindIII under the control of the cytomegalovirus immediate early promoter (Genscript Biotech, USA).

### Epitope prediction and population coverage analysis

Homology modeling was carried out using Modeller tool (v)^56^, 100 models were predicted and the model with the lowest DOPE score was used for the analysis. Energy minimization was carried out using Schrodinger module (v). B-cell discontinuous epitopes were predicted for the PDB templates and the construct sequences using from the DiscoTope webserver (IEDB) using the default parameters^57^. This tool uses solvent-accessible surface area and contact distances for predicting structural B-cell epitopes. T-cell epitopes were predicted using the NetMHCpan EL 4.1 (MHCI) and a combination of NetMHCIIpan 4.0, NN-align 2.3 and SMMalign (MHCII) from IEDB^58,59^. The thresholds used were <0.5 for strong binders and <2 for weak binders for MHCI and <2 for strong binders and <10 for weak binders.

### *In vitro* expression and Immunofluorescence assays

HEK293T cells were transfected with DDV using X-tremeGENE HP DNA transfection reagent as per the manufacturer’s instructions. The transfected cell lysate and supernatants were collected 36 hours of post-transfection, and antigen expression was confirmed by western blot analysis. Cells were washed with phosphate-buffered saline (PBS) and lysed with NP40 supplemented with protease inhibitor cocktail (Roche) and 1mM PMSF(Sigma) was used to make cell lysates. Protein lysates were separated on SDS-polyacrylamide gel, transferred onto a nitrocellulose membrane (Bio-Rad), and blocked for 1 h in 5% skimmed milk. Subsequently, membranes were incubated in mouse antisera (1:200 dilution) against DDV. Secondary antibodies conjugated to horseradish peroxidase (HRP) were used at a dilution of 1:1500. After washing with PBS/PBST the blots are developed using an enhanced chemiluminescence system (Thermo Fisher).

For Immunofluorescence, about 10^5^ cells were plated on a coverslip. The next day, cells were fixed in ice-cold methanol, permeabilized with 0.1% Triton X-100 for 10 min, followed by blocking with 1% BSA for 30 min at room temperature (RT). Permeabilized cells were then incubated with antisera (1:100) for 1 hour (h) at RT, washed thrice with 1X PBS, followed by incubated with goat anti-mouse IgG-AF488 at 37°C for 30 min. Cells were again washed thrice with 1X PBS, mounted (Prolong gold antifade Mountant with DAPI, Invitrogen), air-dried, and visualized using an FV1000 confocal microscope.

### Immunization of mice with DENV DNA vaccine construct with electroporation

To determine the immunogenicity of the DENV DNA vaccine constructs, mice were immunized with 50µg of DNA in a total volume of 50µl of sterile water by a syringe into the anterior tibialis (TA) muscles then electroporated using BTX ECM 830 with 8 square 40-V electric pulses in alternating direction with a time constant of 0.05 s and an inter-pulse interval of 1s2^60,61^. Each group received 2 booster doses at 2-week intervals, and mice were euthanized two weeks following the last immunization.

### IgG and IgG subtype binding antibody titers by indirect ELISA

DENV binding antibody titer was analyzed by indirect ELISA. 96 well plates (Thermo Scientific) were coated with recombinant protein in a coating buffer (0.1M NaHCO_3_) and incubated overnight at 4 □C. The following day, plates were blocked with 3% BSA in PBS for 2 hours at room temperature. Triplicate samples of serially di plasma ranging from 1:100 to 1:5,00,000 were added to the plate and incubated for 2 hours at room temperature or overnight at 4 □C. After washing, secondary anti-IgG (Sigma), anti-IgG1(Invitrogen), and anti-IgG2a (Invitrogen) or IgG2c (Abcam) antibodies conjugated with horseradish peroxidase was added at 1:2000 (IgG, IgG1, IgG2a) and 1:5000 (IgG2c) dilution for 1 hour at room temperature. The plates were then developed with TMB substrate (Sigma) for 15-20 minutes. The reaction was stopped with a stop buffer (Invitrogen) and the optical density (O.D) measured at 450 nm. Cut-off values for each dilution were set using the O.D of naive samples in the formula: naive O.D at a dilution + (2.5 * standard deviation). Starting from the lowest dilution, the sample dilution prior to the one which was exceeded by the cut-off was considered as the end titer value^62,63^.

### Flow cytometry-based neutralization test (FNT)

The flow cytometry-based neutralization assays were performed in triplicate in 96-well cell culture plates with flatbottom wells. Each well contained 5 × 10^4^ DCSIGN-expressing U937 cells. Immune sera were serially diluted, and the virus was pre incubated with the sera for 1 h at 37°C. The cells were washed, and virus and serum mixture was added to the cells for 1 h at 37°C. Next, the wells were filled with cell culture medium, and the plates were incubated for 24 to 48 h at 37°C in 5% CO2. The cells were prepared for flow cytometry analysis by washing them in phosphate-buffered saline and transferring them to 96-well plates with round-bottom wells. The cells were fixed and permeabilized by using a Cytofix/ Cytoperm kit (BD-PharMingen, San Diego, CA) and stained with fluorescein isothiocyanate-conjugated monoclonal antibody 4G2, a monoclonal antibody that recognizes the flavivirus E protein. The cells were analyzed with a FACScan flow cytometer. The serum dilution that neutralized 50% of the viruses was calculated by nonlinear, dose-response regression analysis with Prism 4.0 software (GraphPad Software, Inc., San Diego, CA).

### IFN-γ Enzyme-Linked Immunosorbent Spot (ELISPOT) Assay

ELISpot was performed with the Mouse IFN-γ ELISpot BASIC Kit (Mabtech). In brief, freshly isolated 0.5M splenocytes/animals were plated into polyvinylidene fluoride (PVDF)-coated 96-well plates containing IFN-γ capture antibodies. Cells were stimulated with immunogenic 9-mer peptides predicted from DENV ED III and NS1. Negative control wells contained no peptide. Phorbol myristate acetate (PMA) / Ionomycin (IO) used as a positive control. After overnight stimulation, plates were washed and sequentially incubated with biotinylated IFN-γ detection antibody (R4-6A2), streptavidin-ALP, and finally BCIP/NBT. Plates were imaged with ImmunoSpot Analyzer and quantified with ImmunoSpot software.

### Intracellular cytokine staining

Intracellular cytokine staining was performed by stimulating freshly isolated splenocytes with 50 ng/mL PMA and 500 ng/mL IO in the presence of Brefeldin for 5-6 h. After stimulation, surface staining of CD4, and CD8 was performed, followed by intracellular staining of IFN-γ (Biolegend). Data acquisition was performed on a BD LSRFortessa and analyzed with FlowJo.

### Immune and inflammatory gene expression profile

Mice were sacrificed 2 weeks post-second immunization and lymph nodes were collected for analysis of genes involved in immune response. Lymph nodes were homogenized, and RNA was extracted with TRIzol LS. RNA (50 ng) from whole blood cells and lymph nodes were hybridized to the NanoString nCounter mouse inflammation and immunology v2 panels (NanoString Technologies), respectively. RNA was hybridized with reconstituted CodeSet and ProbeSet. Reactions were incubated for 24 h at 65C and ramped down to 4C. Hybridized samples were then immobilized onto a nCounter cartridge and imaged on a nCounter SPRINT (NanoString Technologies). Data were analyzed with nSolver Analysis software and PRISM. For normalization, samples were excluded when percentage field of vision registration was <75, binding density outside the range 0.1–1.8, positive control R2 value was <0.95, and 0.5 fM positive control was %2 SD above the mean of the negative controls. Background subtraction was performed by subtracting estimated background from the geometric means of the raw counts of negative control probes. Probe counts less than the background were floored to a value of 1. The geometric mean of positive controls was used to compute positive control normalization parameters. Samples with normalization factors outside 0.3–3.0 were excluded. The geometric mean of housekeeping genes was used to compute the reference normalization factor. Samples with reference factors outside the 0.10–10.0 range were also excluded. To identify DEGs between groups, Graph pad Prism software was used to analyze variance with a cutoff of p < 0.05. Log 2 fold changes generated were used for volcano plots constructed with Prism 5 software. DEGs were identified by a fold change cutoff of 1.5. Unsupervised PCA was performed to visualize variability between DDV and pVAX1 control animals.

### Protective efficacy in AG129 mice

AG129 mice [deficient in interferon-α/β and γ receptors] were obtained from B&K Universal (UK). The mice were bred and maintained under specific pathogen-free conditions in individually ventilated cages. 3-4 weeks old AG129 mice were injected intraperitoneal with 100 µl and 300 µl immune serum obtained from vaccinated BALB/c mice. After 2 hrs of passive immunization, the mice were subjected to DENV challenge with 1 × 10^4^ pfu of DENV-2 strain^64^ through subcutaneous route. In pVAX1 control group, the AG129 mice were passively immunized with serum from pVAX1 control group of BALB/c mice. Mice with DENV-2 virus challenge alone were kept as infection control. After infection, each mouse was observed for development of clinical symptoms and body weight changes till mortality or 14^th^ day post-infection. The clinical symptom scoring was done based on the following criteria: Score 1: ruffled fur; Score 2: 1+ hunched back; Score 3: 2+ slow movements or lethargy; Score 4: 3+ facial edema; Score 5: 4+ facial edema with closed eyes; Score 6: 5+hemorrhage (bloody stool) or limb paralysis; Score 7: death. All the experimental procedures were approved by the Institutional Animal Ethics Committee (IAEC) of Rajiv Gandhi Centre for Biotechnology (RGCB) (IAEC/820/SREE/2020).

## Data Availability

Whole Dengue virus genome sequences generated in the study have been deposited in the GenBank. The authors confirm that the data supporting the study findings are available within the manuscript and its supplementary figures and tables. Additional raw data supporting the findings of this study are available from the corresponding authors AS and ES on request.

## Funding

This study was funded by philanthropist Mr. Narayana Murthy, Co-founder of Infosys, NCBS/TIFR, DBT to SK, RGCB (Intramural grant) to ES, and TRC-BIRAC grant to GM. The sponsors of the study had no role in study design, sample collection, data collection, data analysis, data interpretation, or writing of the manuscript.

## Results

### DENV genome sequencing and genotype analysis

In this study, we investigated the genetic diversity of circulating DENV strains in India, designed a DNA vaccine based on circulating DENV genotypes, and evaluated the immunogenicity of this vaccine candidate in two murine models. In essence, we performed a retrospective analysis of 1000 archived serum/plasma samples from NS1 positive patients with dengue clinical presentation collected from 2012–2018 from 4 hospitals in India. Whole-genome sequencing of dengue viruses was carried out from the serum of 319 patients. **Supplementary Table.1** summarizes the sequenced patient’s sample clinical characteristics. Clinical and biochemical correlates of the sequenced samples of this study are consistent with earlier epidemiological observations^65,66^. Of the 319 samples sequenced, we have obtained 120 whole-genome sequences as well as infected serotype infection of 217 patients. Among the 217 patients, 174 patients were infected with a single DENV serotype (mono-infection), and 43 patients had concurrent DENV co-infections. From the 174 patients, 68 (39.1%) of them typed as DENV1, 63 (36.2%) of them as DENV2, 29 (16.7%) of them as DENV3, and 14 (8%) of them as DENV4. The distribution of serotypes during the sample collection period is shown in Figure 1A. Our dengue surveillance data collectively suggest that India is endemic to all four serotypes therefore, an effective vaccine must be tetravalent.

**Figure 1.**
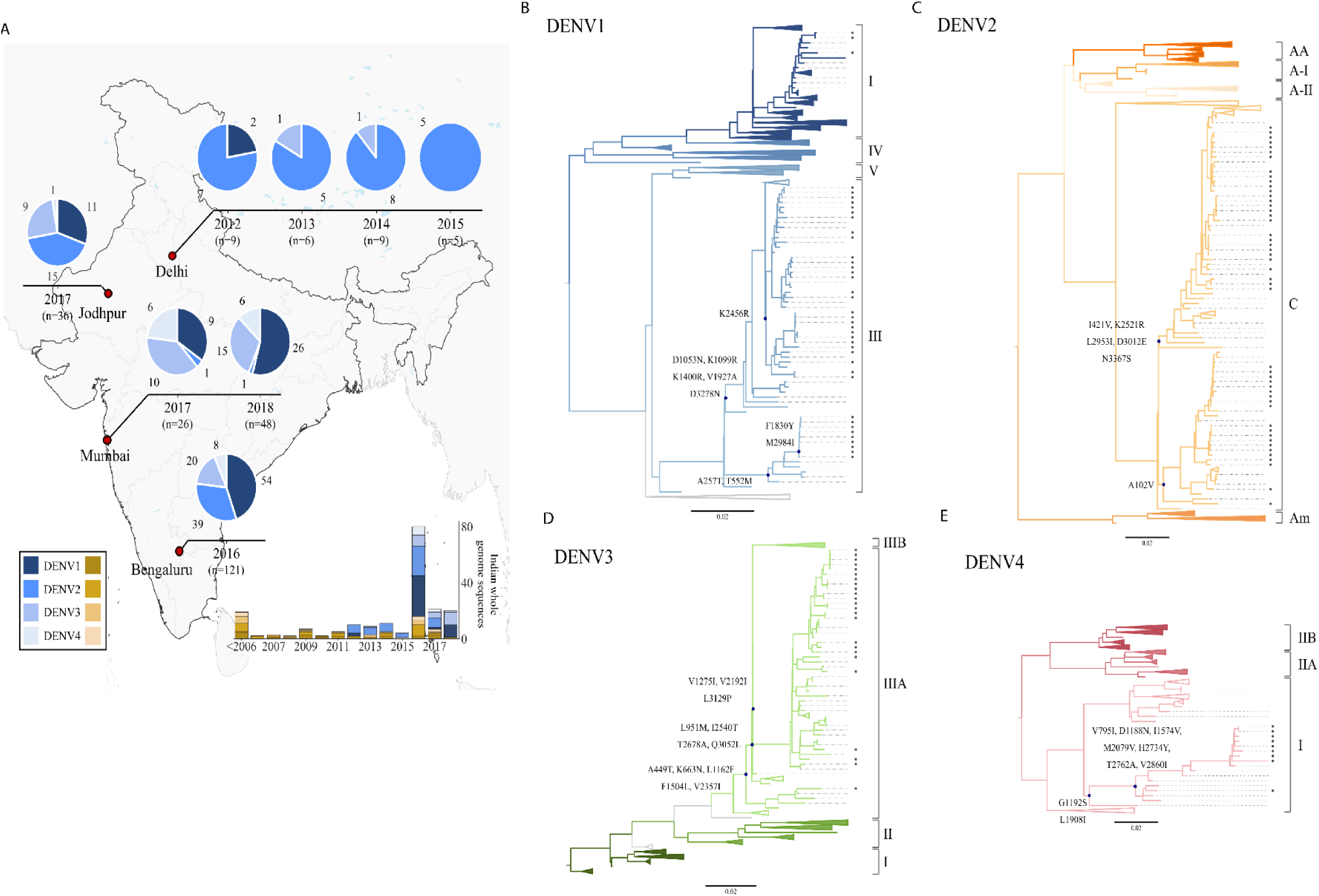
DENV Serotype distribution across sites. **A)** Distribution of DENV serotypes 1-4 from the sites from which samples were received for this study. Serotype distribution for each site is shown as pie charts over time. Total numbers from the site are indicated, each serotype is represented by a different colour. **B-E)** Genotype assignment based on maximum likelihood phylogenetic trees. Phylogenetic trees DENV1-4 using the maximum likelihood method are shown. All available (dated) complete coding nucleotide sequences (DENV1 n = 1800, DENV2 n = 1395, DENV3 n = 823, DENV4 n = 220) from human host, were used for tree construction. Sylvatic strains EF457905 (for DENV1), EF105379 (for DENV2), KT424097 (for DENV3) and JF262779-80 (for DENV4) were used as outgroups to root the tree. Branches have been collapsed to aid visualization. Greyscale colours show different genotypes. Region, where Indian sequences are present, are colored in blue. The detailed tree structure is shown only for the neighbouring sequences. Amino acid mutations with respect to the recent ancestor are shown near important nodes (marked in blue circles).

Furthermore, the phylogenetic-based genotype analysis of our study revealed that all Indian DENV 1-4 strains belong to a single genotype within the serotypes, except DENV1. DENV1, genotype III is the most dominant, apart from a few sequences of genotype I. DENV2 sequences belong to the cosmopolitan genotype, whereas DENV3 and DENV4 sequences belong to genotype III and I, respectively. Our data show that two different DENV2 lineages within the cosmopolitan genotype are propagating simultaneously in all four sites in this study. Both the lineages emerged in the middle of the 1980s. Most of the neighboring sequences are from Asian countries. However, one cluster in the cosmopolitan-b lineage that is found in Kenya and one sequence of a traveler from Ethiopia and Djibouti. Our findings are also consistent with a previous report from Kenya^28^. All the Indian sequences of DENV3 genotype III cluster with other Asian countries. The only genotype I of DENV4 is observed in India. All Indian DENV4-I sequences along with one sequence from Pakistan cluster separately within genotype I from other Southeast Asian sequences. This data highlights that even though there is an enormous intra-serotype variation recorded all over the world, India has distinct genotypes for all four DENV serotypes with prominent intermixing between neighboring countries (Figure 1B-E). Studies performed in India during the previous years also have reported the same^23–25^. Based on the implications of our findings on Indian-specific genotypes, we proposed a vaccine design approach against circulating strains, which may be a better representation rather than strains not found here.

### EDIII global and local genetic diversity and target for vaccine design

The E protein, primarily DIII domain of the E protein, stimulates host immune responses by inducing protective and neutralizing antibodies. Previous studies have demonstrated that mutations in the EDIII region could potentially impact the neutralization of DENV and the host-receptor interaction. Hence, we investigated EDIII protein diversity in global strains and how our study sequences are diverging from international reference strains.

To assess antigenic differences between the EDIII of DENV 1-4 strains from distinct parts of the world, we generated a neighbor-joining tree and selected representative genotype variants from each branch to comprehensively represent global diversity within the serotype. We performed a sequence alignment of these proteins and compared their EDIII genetic diversity relative to the respective reference strains. Variable sites were designated when at least one virus showed an amino acid change at any of the 103 amino acid positions in the alignment. While DENV genotypes are closely related, considerable genetic variation was observed across genotypes of the same serotype in the EDIII region with a range of 1.62-5.83% **(Supplementary Table 2),** which is consistent with an earlier report^26^.

Further, EDIII amino acid sequences of DENV 1-4 samples from our study were compared for their similarity with the wild-type DENV strains. Even though India has its unique genotypes, considerable genetic variations were found in the EDIII region across genotypes of the same serotype. Alignment of the EDIII amino acid sequences revealed 9, 7, 2, and 8 sites of variation in DENV1, DENV2, DENV3, and DENV4 among Indian genotypes, respectively (Figure 2A). Among all four strains, DENV1 exhibited the highest EDIII diversity with a median of 4.85% (range, 2.91%-5.83%), followed by DENV4 and DENV2 with 2.91% (range, 2.91%-3.88%) and 1.94% (range, 0.97 %-3.88%), respectively. The percentage diversity of DENV3 EDIII sequences was found to be limited, ranging from 0% to 0.97 %. The frequency of amino acid variants was calculated across strains and expressed as a percentage (Figure 2 B, D, F **&** H). These mutations were also spotted on the EDIII PDB structure (Figure 2 C, E, G **&** I). Additionally, we also identified several highly variable sites across serotypes. A site was considered highly variable when greater than 50% of the study sequences showed a mutation at that position. The number of highly variable sites in domain III were 5, 3, 2, and 1 in DENV1, DENV4, DENV2, and DENV3 strains, respectively. In addition to our study sequences, we also investigated EDIII mutations in all Indian DENV1-4 strain sequences deposited in the ViPR database. The Supplementary figure 1 shows amino acid variants and their frequency. In line with previous reports, our analysis implicates the EDIII amino acid residues as sites under immune pressure^67^.

**Figure 2.**
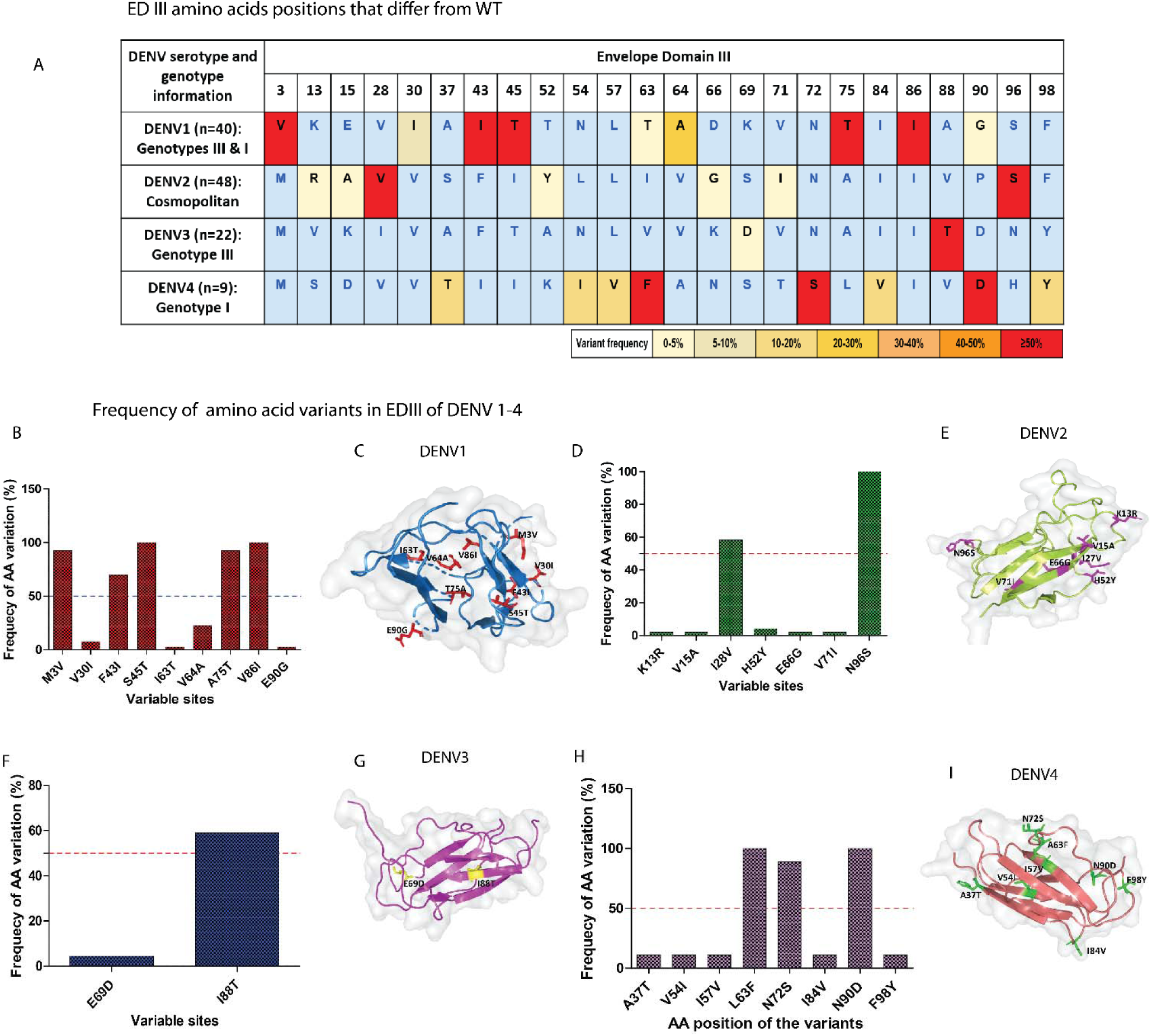
DENV amino acid variation in our study sequences for all four serotypes. **A)** Frequency based representation of all the variable sites, for each serotype within our study sequences. Colours assigned based on the frequency of sequences bearing a mutation at that particular site. **B, D, F and H)** Frequencies of variants within DENV1, DENV2, DENV3, DENV4 strains of our study sequences, respectively. Amino acid variants residue identified relative to (NC_001477(DENV1), NC_001474 (DENV2), NC_001475 (DENV3), NC_002640 (DENV4). **C, E, G and I**) Amino acid variants also spotted on EDIII PDB structure (ribbon). Stick and ball representation on PDB structure indicates variants at the particular position. PDB: 4gt0.1, 4ut6.2.8,4Gsx.1 and 5BIC.1 used to annotate the genetic variants of DENV.

### DENV DNA vaccine construction

To reduce the genetic diversity within serotypes, we selected the best representative Indian contemporary strains-based consensus sequence for vaccine development. We have generated consensus DENV EDIII and NS1 protein sequences from the whole genome sequences obtained from our study (DENV1 n =40, DENV2 n =48, DENV3 n =22, DENV4 n =9) and India-specific DENV sequences (from 2000-2019) that were retrieved from ViPR database. Total number of sequences used to create consensus sequences: (DENV1 n = 142, DENV2 n = 252, DENV3 n = 69, DENV4 n = 52). The Aliview and Geneious tools were used to align and select the consensus amino acid sequence. A consensus was generated from the most frequent residues (bases matching at least 65% of sequences) at each site. Sequences were codon and RNA optimized, synthesized commercially, cloned into pVAX1 expression vector and the generated plasmid was designated as DDV (Dengue DNA Vaccine) (Figure 3A **&** B).

**Figure 3.**
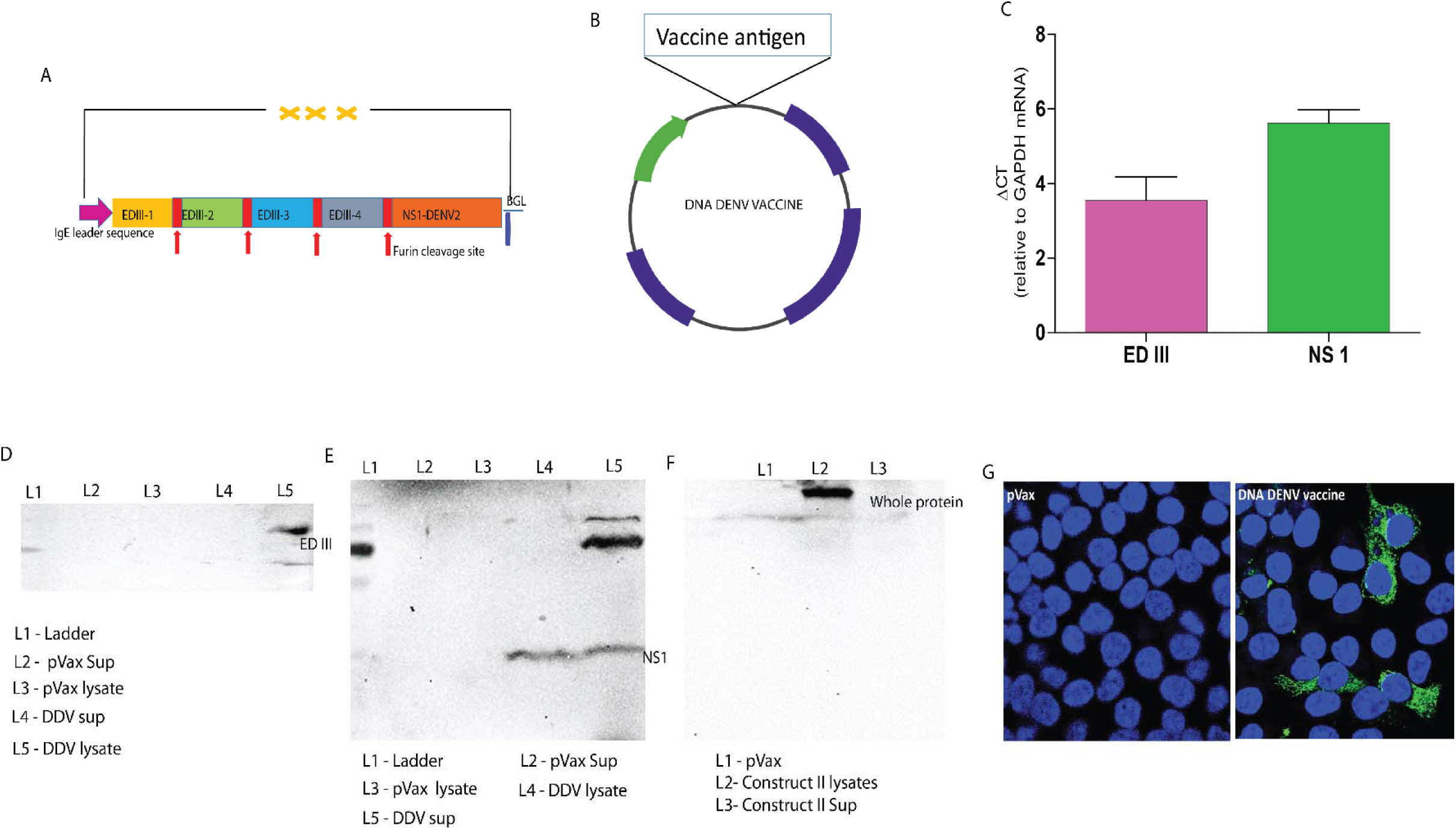
DENV DNA vaccine construction. **A & B)** Schematic representation of DENV DNA vaccine and cloning strategy. Synthetic DENV expression cassette was inserted into the pVAX1 expression vector in between NheI and HindIII under the control of the cytomegalovirus immediate early promoter. **C)** RT-PCR of RNA extracts from HEK293T cells transfected with DNA DENV vaccine. GAPDH used as an internal expression normalization gene. **D-F)** Analysis of in vitro expression of ED III and NS1 protein after transfection of 293T cells with DDV or plasmid control by Western blot. 293T cells supernatant and lysates resolved on a gel and probed with anti-DDV immune sera. Blots were stripped then probed with β-actin loading control. **G)** Immunofluorescence staining of 293T cells transfected with 5ug/well of DDV or plasmid control. Expression of antigen was measured using anti-DDV immune sera. Cell nuclei were counterstained with DAPI.

It is also noteworthy that approximately 26-50% of Indian DENV 1-4 strains exhibited 100% identity with EDIII sequences represented in DDV. Furthermore, the remaining Indian sequences, for all the serotypes, exhibited greater than 93% identity **(Supplementary Table 3)**. DDV also shares >95.15 identities with the EDIII of the top 1000 international dengue sequences of the cognate serotype in the ViPR database.

### Epitope analysis for the EDIII construct

We predicted the structural stability of the EDIII constructs and checked for the 3D structural conservation at the predicted B-cell discontinuous epitope regions (**Supplementary Table 4**). The homology models for the EDIII constructs were subjected to energy minimization, rmsd (root mean square deviation with the template used for modelling) calculation with the PDB structures and Ramachandran map^68^ (https://saves.mbi.ucla.edu) and energy analysis^69^ (https://prosa.services.came.sbg.ac.at/prosa.php) which predicted that the constructs are stable structurally and energetically (Supplementary Figure 2).

In order to estimate the population coverage of the vaccine constructs, we also predicted the T-cell epitopes and the HLA subtypes predicted to bind to each of the epitopes. This analysis revealed that 90-98% of the world population could recognize the MHCI epitopes (using predicted strong binding epitopes) and 90-99% of the population can recognize the MHCII epitopes (using both strong and weak binding epitopes) (**Supplementary Tables 5-7**).

### *In vitro* antigen expression and localization

We first assessed encoded DENV ED III and NS1 transgene expression at the RNA level in HEK293T cells transfected with DDV. Using the total RNA isolated from the transfected 293T cells, we confirmed the EDIII and NS1 mRNA expression by qRT-PCR (Figure 3C). *In vitro,* EDIII and NS1 protein expression in HEK-293T cells was measured by western blot using anti-DDV immune sera on cell lysates. Western blots of the lysates of HEK-293T cells transfected with DDV construct revealed bands near predicted molecular weights of ≈11 kDa (Figure 3D) and ≈48kDa for ED III and NS1 (Figure 3E), respectively. We also detected secreted DENV NS1 in the culture supernatants of transfected HEK-293T cells. EDIII and NS1 protein expression were further validated using EDIII and NS1 monoclonal antibodies (Supplementary Figure 3). In immunofluorescence studies, the ED III and NS1 protein was detected in HEK-293T cells transfected with DDV and exhibit antigen staining of the expressed proteins mainly in the cytoplasm which suggested the immune reactivity of the encoded protein (Figure 3G). In summary, *in vitro* studies revealed the expression of antigens at both the RNA and protein levels after transfection of cell lines with the candidate vaccine construct DDV.

### Induction of humoral immune responses against DDV in BALB/c and C57BL/6J

DENV specific humoral responses following vaccination were characterized in two different murine strains, BALB/c and C57BL/6J. Mice (n=6) were vaccinated three times, two weeks apart, with 50 ug of the DNA vaccines or control pVAX11 plasmid vector using Tabialis anterior (TA) muscle delivery. Vaccinated mice were bled at day 0 and two weeks after each vaccination to obtain sera, which were assayed for the presence of DENV antibodies by enzyme-linked immunosorbent assay (ELISA) against recombinant protein as a capture protein (Figure 4A). Binding antibody ELISA data revealed that the DDV induced DENV specific antibody responses. The results showed that all mice developed anti-DENV antibodies after a single immunization. The anti-dengue IgG responses were significantly increased after 1-2 booster immunizations, appearing to peak 2 weeks after the second booster in both BALB/c and C57BL/6J strains (Figure 4B **&** D). Comparison between the serum IgG endpoint titers of DDV and plasmid control groups in both murine models showed robust titers elicited by DDV immunization. The endpoint tiers ranged from 1:1000 to 1:500000 in individual animals (Figure 4 C **&** E). These findings demonstrated the ability of the DENV DNA vaccine construct to express in mammalian cells potentially and the antibodies induced by these constructs were able to react with the Dengue vaccine antigens.

**Figure 4.**
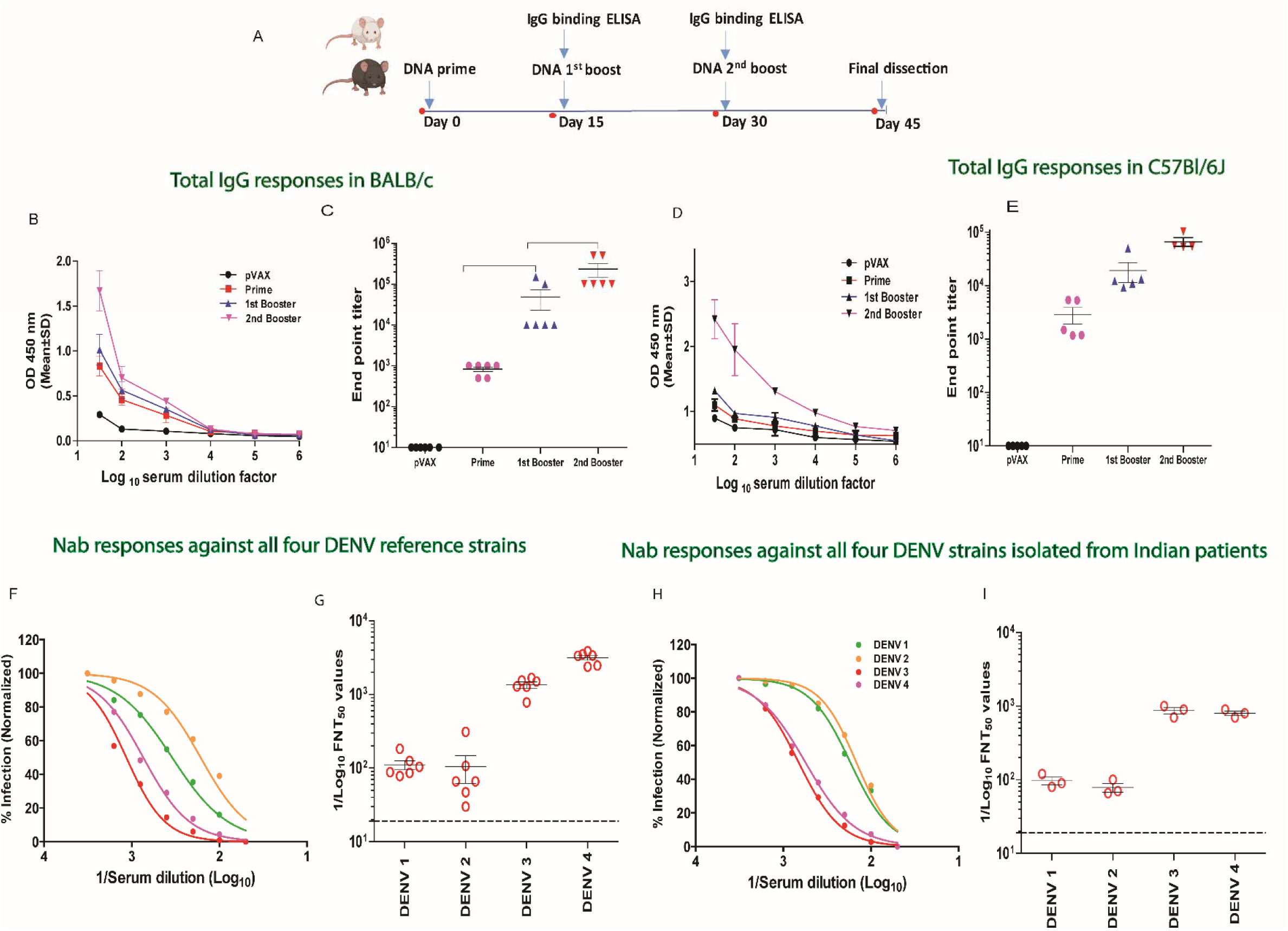
Antibody response induced by DENV DNA vaccination in BALB/c and C57BL/6J mice. **A)** Schedule of vaccination and antibody assays. BALB/c and C57BL/6J mice were immunized by TA injection of 50ug DDV or plasmid vector at day 0,15 and 30. Sera were collected at each time points. **B-E)** Indirect ELISA reactivity showing O.D. measured at 450nm (in pooled sera) and Serum IgG binding end point titers (in individual animals) of BALB/c **(B & C)** and C57BL/6J strains **(D & E)**. Antibody titers were measured by ELISA plates coated with purified recombinant DENV DNA vaccine antigens. **F-I)** Sera neutralization titers against DENV 1-4 laboratory prototype strains and recent clinical isolates. Sera was collected at 14 days after second booster dose and analysed for neutralization of DENV by FNT assay. **F & H)** Representative normalized percentage of infection curves are shown from laboratory prototype strains and recent clinical isolates, respectively. FNT50 values were calculated for individual animals and presented in **G & I** from prototype strains and clinical isolates, respectively. Each point represents an individual animal, while horizontal lines indicate the mean, SD.

Flow-based virus neutralization assay was performed in U937-DC-SIGN cells to assess the levels of anti-DDV immune sera-induced neutralizing antibody titers against laboratory DENV strains. DDV-vaccinated mice immune sera showed a clear neutralizing antibody titer against all four serotypes simultaneously; the median FNT50 titers against DENV 1-4 ranged from 182-3500 (Figure 4 F **&** G). As per WHO recommendation for DENV vaccines^70^, we next investigated the neutralizing potency of anti-DDV immune sera against DENV 1-4 recent clinical isolates. DENV 1-4 serotypes were isolated from DENV NS1 positive patients in India and FNT was performed. Our data showed anti-DDV immune sera effectively neutralized Indian isolates as well and FNT50 titers against DENV 1-4 ranged from 150-900 (Figure 4 H **&** I). These data provide evidence that DDV-induced neutralizing antibody responses cover major currently circulating strains in India as well.

### Robust T cell responses induced by DDV

DENV specific cellular responses were assessed in vaccinated animals by ELISPOT. A total of 95 and 344 ED III and NS1 specific T cell peptides, respectively, were identified in the DENV DNA vaccine construct. MHC class I binding predictions, peptide selection, and antigenicity of those peptides were analyzed by NetCTL 1.2 and Vaxijen 2.0. A total of 15 CTL epitopes (9 mer peptides) were screened which 11 were found to possess antigenicity and were chosen for synthesis (Figure 5B). We assayed T cell response against dengue antigens via IFN-γ ELISPOT as the IFN response has previously been shown to be associated with vaccine immunogenicity following yellow fever vaccination^71^. In addition, IFN-γ has been described as a mediator of T cell responses and plays a distinctive role in antiviral activity against dengue viruses^72,73^. Mice were immunized as before. Two weeks after the second booster DDV or pVAX1 plasmid control immunized mice were euthanized and splenocytes were isolated (Figure 5A). Single-cell suspensions were stimulated with the peptide pools (Pool 1- EDIII peptide mixture; Pool 2- NS1 peptide mixture; Pool 3- EDIII and NS1 peptide mixture) number of IFN-γ producing CD8+ T cells was analyzed. Results show that both ED III- and NS1 specific cellular responses (presented as IFN-γ SFU/cells) were detected by ELISPOT in DDV vaccinated animals while negligible spots were detected in the plasmid vector-vaccinated animals (Figure 5 C-F). PMA/IONOMYCIN was used as a non-specific positive control. As expected, stimulation with PMA/IONO in all ELISPOT assays performed with cells from DENV vaccinated or control animals induced a high IFN-γ response.

**Figure 5.**
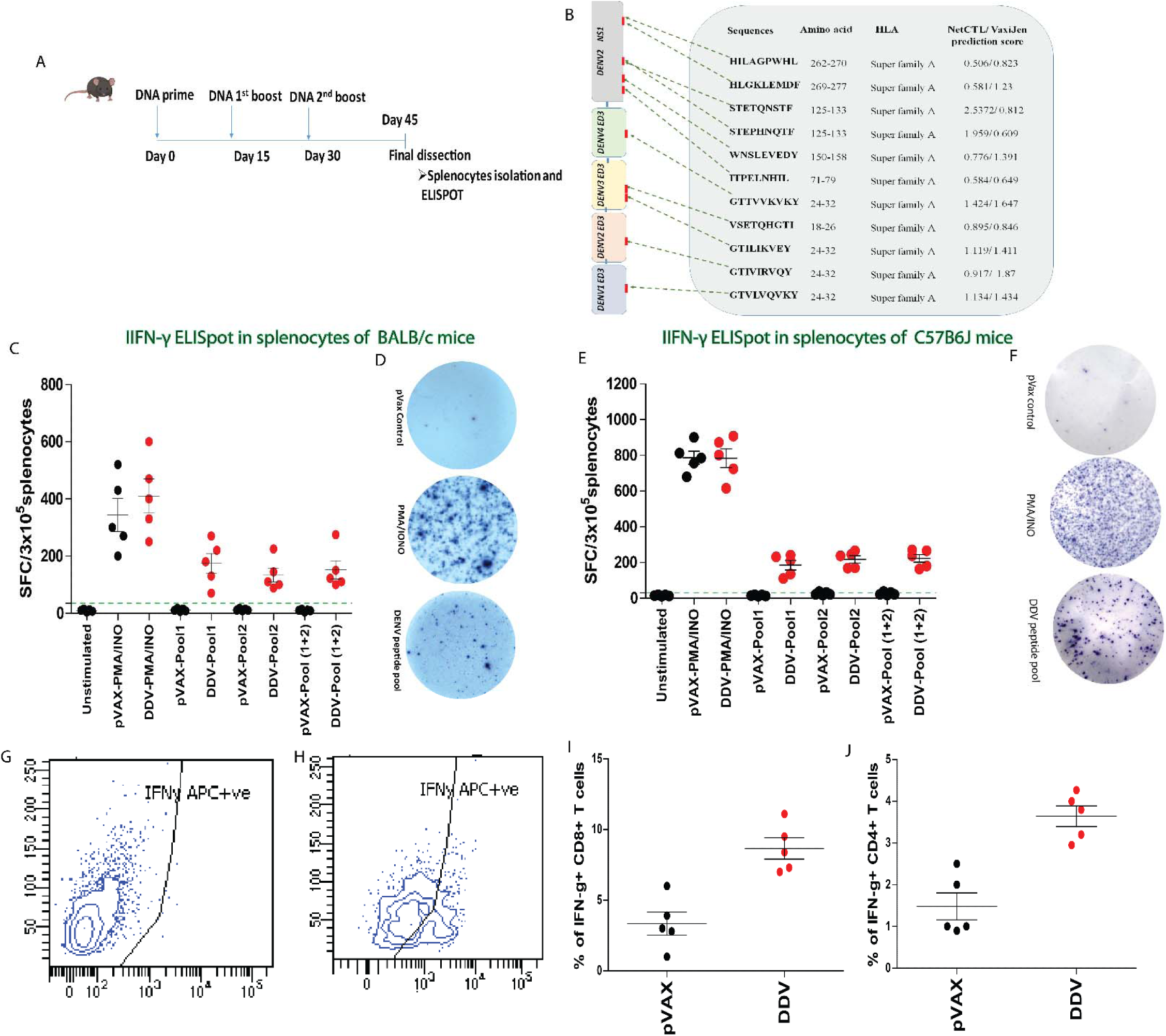
Characterization of cellular immune response induced by DENV DNA vaccine in mice. **A)** Schedule of vaccination and T cell assays. Mice (n□=□6/group) were immunized with 50 µg DENV DNA vaccine. T cell responses were analysed 2 weeks after second booster dose. **B)** Map of the DDV and predicted potential immunodominant peptides through NETCTL and VAXIJEN. **C – F)** Antigen specific T responses to pooled EDIII-NS1 peptides were measured by IFN-γ ELISpot assays after vaccination with DDV or plasmid control in BALB/c and C57BL/6J animals. Bars represent the mean□+□SD. PMA/IONOMYCIN was used as a non-specific positive control. **G-J)** Flow cytometric analysis of Intracellular cytokine staining for IFN-g in C57BL/6J mice splenocytes. **G & H)** Representative image of Intracellular cytokine staining in CD8+ T cells in DDV or plasmid control groups. **I & J)** Percentage of IFNγ+ CD8+ and CD4+ T cells in DDV and plasmid control vaccinated animals determined through ICC.

We further analyzed intracellular IFN-γ in both CD8+ and CD4+ cells in both vaccine and control groups of animals. As a positive stimulus for T cell activation, PMA/Ionomycin were used. Exocytosis of cytokines was blocked by the addition of Brefeldin A (10 ug/ml) during stimulation. Cells were permeabilized, labeled, and fixed for flow cytometry. IFN-γ CD8+ T cells and IFN-γ CD4+ T cells were proportionately higher, as found using intracellular staining with flow cytometry, in DDV as compared to pVAX1 vaccinated animals (Figure 5 I**&**J). In summary, DENV DNA vaccine candidate DDV induces robust T-cell response in mice.

### DDV vaccination skewed a Th1-dominant response

Th1 skewness has been shown to elicit a robust adaptive response in terms of cellular activation and antibody production. The induction of polyfunctional Th1 cells is an essential element of a protective vaccine response^74,75^. Respiratory disease viruses such as SARS-CoV and MERS-CoV vaccine development have highlighted the importance of Th1 skewed response in mitigating the risk of vaccine-induced disease enhancement^75,76^. Thus, the Th1/Th2 balance elicited by vaccination with DENV DNA vaccine was investigated. The IgG subclass fate of plasma cells is highly governed by Th cells. To determine whether DDV showed skewing of Th1 over Th2 responses, measured Th1-associated IgG subclasses-IgG2a (BAlB/c) (Figure 6 B**&**C) and IgG2c (C57BL/6J) (Figure 6 D**&**E)- against the Th2 associated IgG1. C57BL/6J and BALB/c mice are prototypical Th1 and Th2 animal strains, with C57BL/6J producing high IgG2c and BALB/c, mostly IgG2a. Hence, in C57BL/6J mice, evaluation of IgG2c and IFN-γ is critical to correct interpretation of Th1 immune responses. In both strains of mice, DDV vaccination-induced Th1 skewed IgG subclass responses. These findings indicate that the DDV elicits Th1 dominant immune responses.

**Figure 6.**
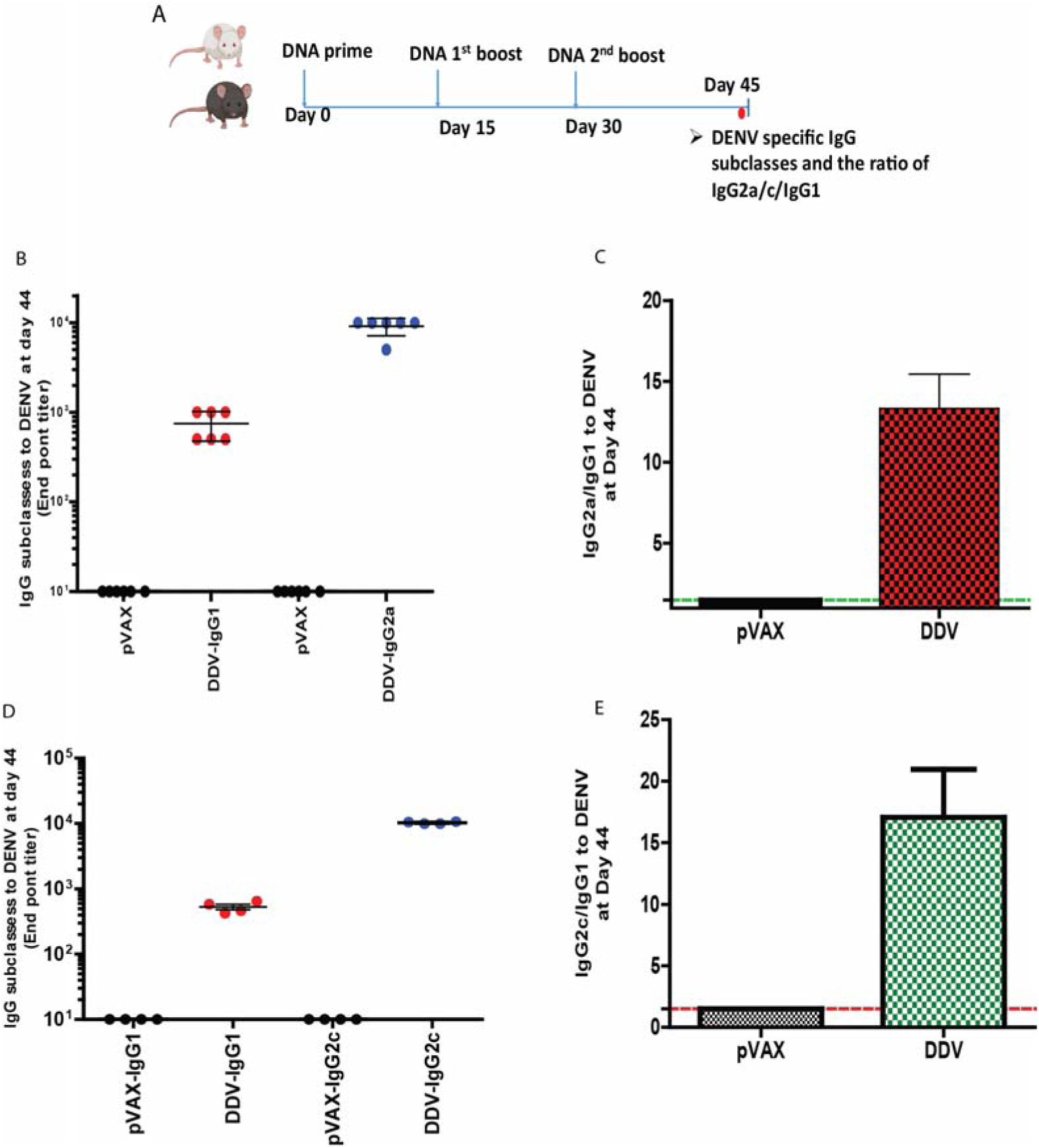
DDV elicits Th1-biased immune responses. **A)** Th1/Th2 Assay schedule: BALB/c and C57BL/6J mice were immunized by TA injection of 50ug DDV or pVAX1 at day 0,15 and 30. two weeks after the 2^nd^ dose sera collected and assayed for IgG subclass antibodies. **B & C)** DENV specific IgG subclasses and the ratio of IgG2a/IgG1 in BALB/c mice immunized with DDV or plasmid control. **D & E)** DENV specific IgG subclasses and the ratio of IgG2c/IgG1 in C57BL/6J mice immunized with DDV or plasmid control.

### Immune gene expression following DDV vaccination

To determine how the DDV exerts its immunogenicity, C57BL/6J mice were vaccinated thrice, two weeks apart, with 50 ug of the DDV and compared these to equivalent doses of pVAX1 vector control. Since immune responses develop in germinal centers in draining lymph nodes, the inguinal lymph nodes of mice at two weeks after 2^nd^ booster were isolated and analyzed with the Nanostring V2 immunology panel (Figure 7A). Principal-component analysis (PCA) of immune gene expression showed clustering of responses to DDV distinct from pVAX1 plasmid controls, indicating clear differences in immune gene expression following DDV vaccination (Figure 7B).

**Figure 7.**
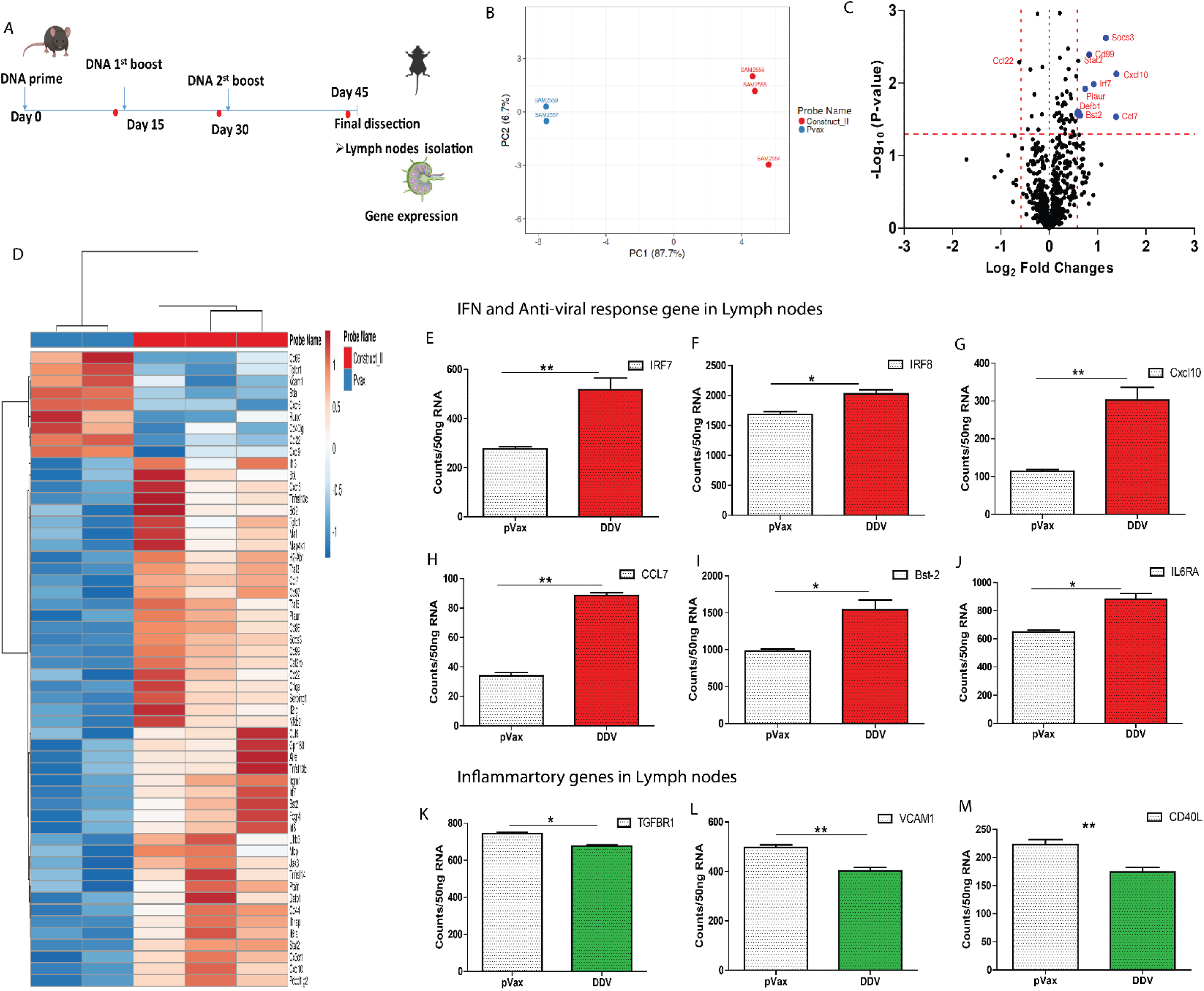
Transcriptomic analysis of immune genes after vaccination with DDV. **A)** C57BL/6J mice were immunized with either pVAX1 or DDV and animals were sacrificed at 14 days after 2^nd^ Booster dose and lymph nodes were harvested. Gene expression of immune genes were measured in lymph nodes. **B)** Principal-component analysis (PCA) of immune gene expression following vaccination with DDV or pVAX1 control. **C)** Volcano plots of fold change of DDV versus pVAX1 control (x axis) and log 10 p value of DDV versus pVAX1 control (y axis). A deviation of 1.5 log2 fold change in gene expression was set as the cut-off value. **D)** Differentially expressed genes DDV and plasmid control in lymph nodes presented as heatmap of Z scores. **E-M)** Lymph nodes transcriptomic data at 14 days after 2^nd^ booster immunization showing Nanostring counts per 50ng RNA of selected IFN and inflammatory genes.

Differentially expressed genes were examined in the lymph nodes of mice injected with DDV compared with those administered a similar dose of pVAX1 plasmid control (Figure 7D). Volcano plot analysis identified significant enrichment of various innate and adaptive immune responses, lymphocyte activation, cytokine, interferon signaling, and Class I antigen presentation genes in DNA vaccine immunized animals (Figure 7C). Some of the most highly expressed genes included Cxcl10, Socs3, Ccl7, Plaur and Defb1 which play roles in directing Th1 and Th2 effector responses. These genes have also been linked to antigen presentation, innate and adaptive immune cell recruitment, T cell stimulation, and dendritic cell maturation in the context of host immunity^77–82^. Furthermore, there are several antiviral defense and IFN-Th1 responsive genes that were also activated upon DENV DNA vaccination. These include Stat2 (IFN signaling), Irf7 (IFN inducible genes expressed on Th1 cells), Bst2 (development of antiviral T cell distribution and function in addition to augmenting DC activation), and CD 99 ( Th1 type cytokine response)^83–85^ (Figure 7 E-J). These findings collectively suggest the development of the immune response in the inguinal lymph nodes of mice immunized with DDV.

### Passive transfer of serum from DENV DNA vaccinated animals protects against lethal DENV2 challenge

To assess the role of humoral immune response in mediating protection from DENV challenge, we passively transferred serum from BALB/c mice immunized with either plasmid control or DDV into AG129 mice. Groups of AG129 mice received anti-pVAX1 sera at 300 µl per mouse or anti-DDV immune sera, at two dosage levels, 100 and 300 µl mice were challenged 2 hours after-passive transfer with a lethal dose of DENV-2 (10^5^ FIU/mouse). The control group did not receive immune sera but were subjected to the lethal challenge dose. All groups were tracked for body weight changes, clinical signs, and survival for up to 14 day’s post-challenge (Figure 8A).

**Figure 8.**
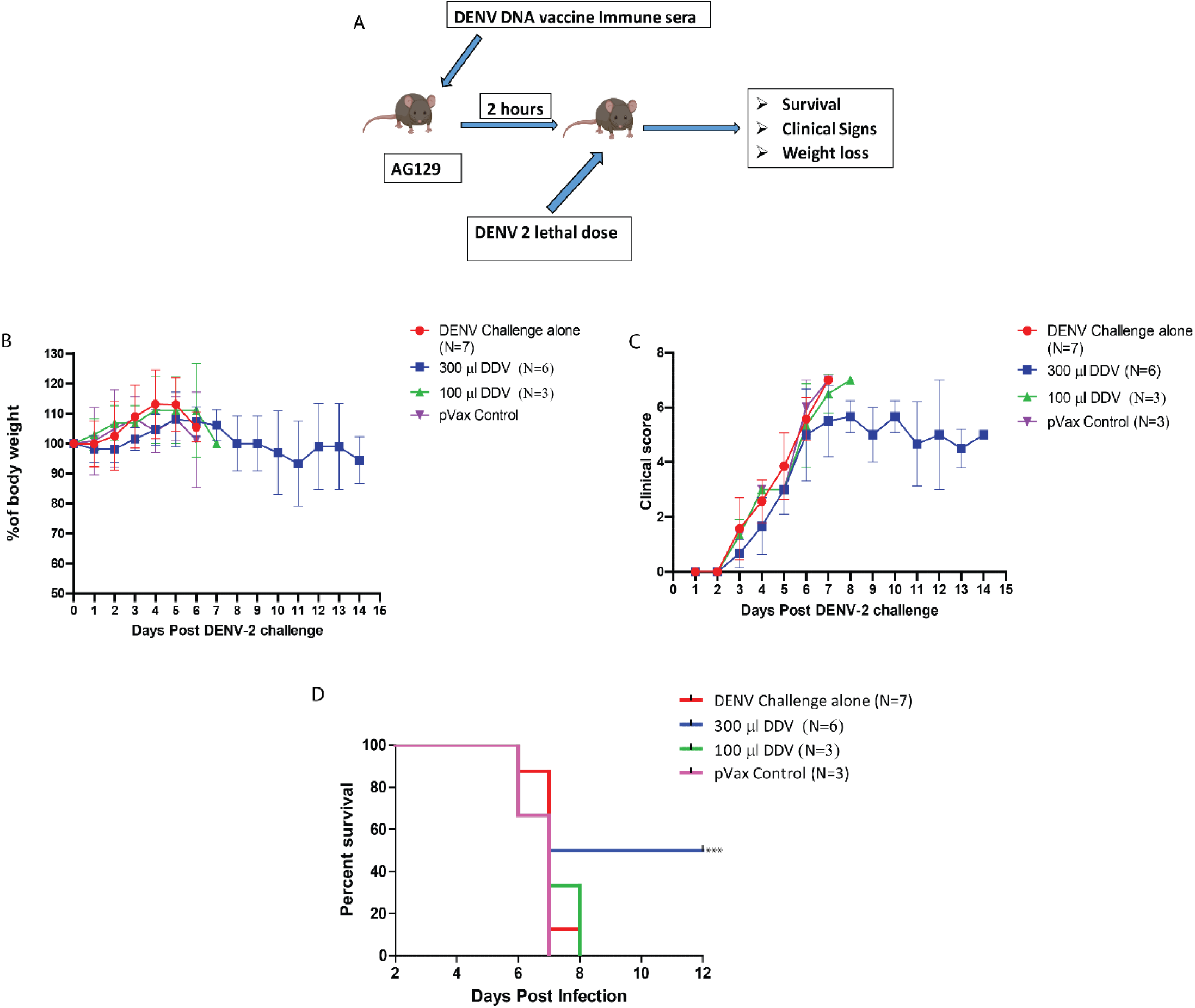
The capacity of anti-DDV immune sera to confer protection against lethal DENV-2 challenge in AG129 mice. **A)** Schematic representation of the experimental design. Groups of AG129 mice were administered (i.p) BALB/c immune sera in two dosage levels, 100 µl and 300 µl/mouse. Two hours after passive transfer the mice were challenged with a lethal dose of DENV 2 (105 FIU/mouse). All groups were monitored for body weight changes **(B),** clinical symptoms **(C)** and Survival **(D).**

The DENV-infection-only group showed an initial increase in body weight; however, with a steep decrease from day 5 or 6. The maintenance of body weight or percentage increase were observed to be better in 300µl immunized mice compared to DENV-challenged group and pVAX1 control group (Figure 8B). The mice started showing noticeable symptoms from 3^rd^ day post-infection starting with ruffled fur which continued to become more aggressive and prominent with the disease progression. Despite the initial rise in symptoms, clinical scoring showed reduced manifestation of the disease in the 300µl immunized group at later stages compared to the infection control and pVAX1 control. However, the 100µl sera immunized group did not show any significant difference in clinical symptoms from both the DENV-challenge and pVAX1 controls (Figure 8C). In the infection control group, the mortality starts on 6^th^ day post-infection and a 100 % mortality was observed by 8^th^ day post-infection (Figure 5D). The pVAX1 control group showed a similar mortality pattern as the infection control group. On the other hand, in the 300µl sera injected group, three mice survived for at least 12-days post-infection out of the total six. The data from the survival curve indicate about a 50% better survival for the 300µl sera vaccinated group compared to the controls. Immunization with 100µl immune sera failed to provide any advantage in survivability compared to the infection and pVAX1 control group.

## Discussion

Dengue vaccine development is complicated by serotype and its intra-serotype diversity. The high mutation rate associated with the RNA viruses leads to different DENV genotypes and further into lineages and clades^86^. The genetic and antigenic differences between genotypes of the same serotype were not considered to impact long-term protective immunity and vaccine efficacy. Several recent studies challenge this assumption. DENV homotypic reinfection, reported in people in Nicaragua and Peru, is potentially driven by genotype differences between primary and secondary infecting viruses^17,18,87^. Some studies have demonstrated that antigenic variation of genotypes can have a high impact on the breadth of antibody neutralization against different genotypes elicited by immune sera from people who have been exposed to natural infections or vaccination^88^. Petty et al. and Shrestha et.al., demonstrated that monoclonal antibody panels generated against DENV1 genotype II and DENV2 Southeast Asia genotypes have lower neutralizing efficacy against heterotypic genotypes of the same serotypes^89,90^. A similar study on genotype cross-reactivity for DENV3 revealed that DENV3 genotype-I neutralizing antibodies did not confer effective protection against DENV3 genotypes-IV^91^. Moreover, the limited efficacy of Dengvaxia against DENV2 points towards differences between the vaccine strain and disease-causing DENV in the clinical sites^31^. These findings suggest that while selecting vaccine strains, investigators and vaccine developers should consider circulating genotype variants.

Here, we used the whole-genome sequencing approach to investigate the sequence diversity of Indian dengue strains and their distribution patterns. Our sequencing efforts have increased the whole genome sequences from India by 187%. The serotype distribution data of this study revealed a parallel evolution of India-specific genotypes for all four serotypes of Dengue virus by genetic drift with continuous exchange among various regions. Despite globalization and frequent traveling, genotypes are contained within the geographical boundaries with prominent intermixing between neighboring countries. Since the genotypes are bounded geographically, the vaccines should be tested at different locations before approval, or they can be restricted to areas having similar genotypes.

EDIII is a potent immunogen known to elicit serotype-specific antibodies and is less likely to enhance dengue viral infection^34,36^. Mutations in EDIII of DENV have been reported to influence the neutralization resulting in escape variants^92^. Previous studies have indicated that DENV EDIII region has the highest sequence heterogeneity among genotypes than Domain I or II of the envelope protein^67^. Yang et al. and Wahala et. al. also demonstrated that DENV3 genotype-specific EDIII mutations in the type-specific mAbs (8Ab and 1H9) binding sites lead to significant variation or loss of neutralization efficacy across DENV3 genotypes^93,94^. Sequencing data of our study revealed that there were multiple mutations in EDIII region, both within and across genotypes of all four serotypes circulating in this region. These mutations could impact the neutralization of viruses and need further investigation. Furthermore, the likelihood of cross-protection across genotypes following vaccination is also debatable.

Here we developed a polyvalent DENV vaccine construct encoding synthetic consensus EDIII of DENV 1-4 and DENV2-NS1 and delivered using DNA plasmid. Despite promising results in preclinical trials, the low immunogenicity of DNA vaccines limits their development for human use^95^. In this study, DDV was subjected to codon optimization, RNA optimization, and IgE leader sequence utilization which was reported to be favorable to increase the immunogenicity in CHIKV^96^, Zika^97^, MERS^98^, and SARS-CoV2^8,99,100^. DNA vaccines. In proof-of-principle studies, we showed that vaccination of mice with DDV delivered *in vivo* by intramuscular EP induced robust anti-DENV reactive IgG responses in both BALB/c and C57BL/6J strains. Importantly, antibody response induced by the DNA vaccine broadly neutralized the infectivity of local and global DENV 1-4 serotypes belonging to the different genotypes. The data indicates that the polyclonal repertoire of antibodies induced by DNA vaccine has adequate breadth of antigenic specificity encompassing multiple strains within each of the four serotypes.

A neutralizing antibody response is an established correlate of protection following vaccination against several flaviviruses including, ZIKA and yellow fever virus^101–103^, and likely contributes to protection following immunization with DDV. Our data indicate that passive transfer of serum from DDV immunized mice was sufficient to confer protection against lethal DENV2 challenge in mice. Our data is also comparable with EDIII VLP based tetravalent DENV vaccine^36^. Critically, it was also observed that passive transfer of immune sera reduces clinical severity; it could be due to the NS1 immunity and B cell memory induced by the vaccine candidate. Moreover, the study substantiates DDV-mediated protection from a lethal DENV2 viral challenge despite the low neutralizing antibody titers produced. To our knowledge, there is no epidemiological data on the magnitude of the neutralizing antibody titers necessary for protection against DENV infection. This data supports the quality of EDIII specific antibodies, which could protect against DENV infection.

Although dengue neutralizing antibodies are capable of protecting against dengue infection, there is growing consensus that the optimal tetravalent dengue vaccine should generate neutralizing antibodies as well as T-cell responses^104^. Antigen-specific T cell responses live longer than neutralizing antibodies and provide durable protection, making them critical for DENV vaccine development^105^. DDV was found to be capable of inducing antigen-specific T cell responses in mice, as evidenced by the generation of IFN-γ in vaccinated animal’s splenocytes. It is well known that IFN signaling plays a key role in priming adaptive T cell responses and directly influences the fate of both CD4+ and CD8+ T cells during the initial phases of antigen presentation, thus shaping the effector and memory T cell pool^106–108^. The production of IFN-γ cytokine has previously been shown to be associated with protection against DENV in experimental animals as well as humans^109^. Overall, our data indicate that antigen-specific T cell immunity is indeed robustly activated following immunization of DDV. Furthermore, like whole virus-based vaccines and viral vector vaccines, DDV elicits robust humoral and cellular immune responses, demonstrating the advantage of DNA vaccines.

Our results indicated that several innate and adaptive immune genes are induced by DDV and can be used to predict the strength of the immune response. In terms of adaptive immunity, DDV was able to elicit DENV specific Th1-predominant responses, as revealed by the IgG(2a/2c)/ IgG1 subtype ratio of antibodies induced by DENV DNA vaccine. Th1 skewness has been shown to elicit robust adaptive responses in terms of cellular activation and antibody production, while Th2 cells are largely responsible for generating the humoral response^74^. Furthermore, Th1 memory cells, especially CD8+ populations, confer long-term protection through rapid expansion and viral clearance^110^. One important advantage of using DNA EDIII-NS1 -based vaccine candidate is that DDV-induced predominantly a Th1 biased T cell response, thereby reducing the risk of potential ADE. Additionally, immunological imbalance and exacerbated disease pathology due to Th2 skewness highlight the necessity of Th1 biased immunity in vaccination regimens^110,111^.

In summary, DENV genome surveillance confirms the serotypes and their genotype diversity in India. We describe the generation of a DENV vaccine based on circulating genotype sequence information, utilizing optimized synthetic DNA plasmid for delivery. Our *In silico* structural analysis of EDIII consensus vaccine sequence revealed that epitopes are structurally conserved and immunogenic across HLA diversity. We have shown that immunization with DENV DNA vaccine resulted in robust neutralizing antibody responses against all four serotypes simultaneously and passive transfer of vaccinated immune sera confer protection against lethal DENV challenge. In addition, our DNA vaccine candidate induces multifunctional antigen-specific T cell responses. Taken together, DDV is a promising vaccine candidate that warrants further investigation. Further, our study emphasizes the utility of consensus sequence approach, in conjunction with DNA delivery, for the development of vaccines against various emerging/re-emerging viruses.

## Acknowledgements

The authors thank Dr. Krishnamurthy, Dr. Yogesh Chandra, and Dr. Swetha Reddy for their technical assistance. We acknowledge the staff of the BLiSC Animal Care and Resource Centre and TheraIndx Lifesciences Pvt. Ltd. for assistance with animal husbandry and immunizations. We also thank Rajiv Gandhi Center for Biotechnology (RGCB) central animal facility for carrying out AG129 mice challenge studies. The authors are grateful to the Bioassay lab at THSTI and TheraCUES Innovations Pvt. Ltd. for performing antibody neutralization and nanostring assays, respectively. We would like to thank the BLiSC Central Imaging and Flow Cytometry Facility for help with FACS data curation and image acquisition and the BLiSC Mass spectrometry facility for protein sequence validation. We would also like to thank the C-CAMP sequencing facility and Genotypic India Pvt. Ltd. for sequencing dengue viruses used in this study.

---

**Supplementary figure 1.**
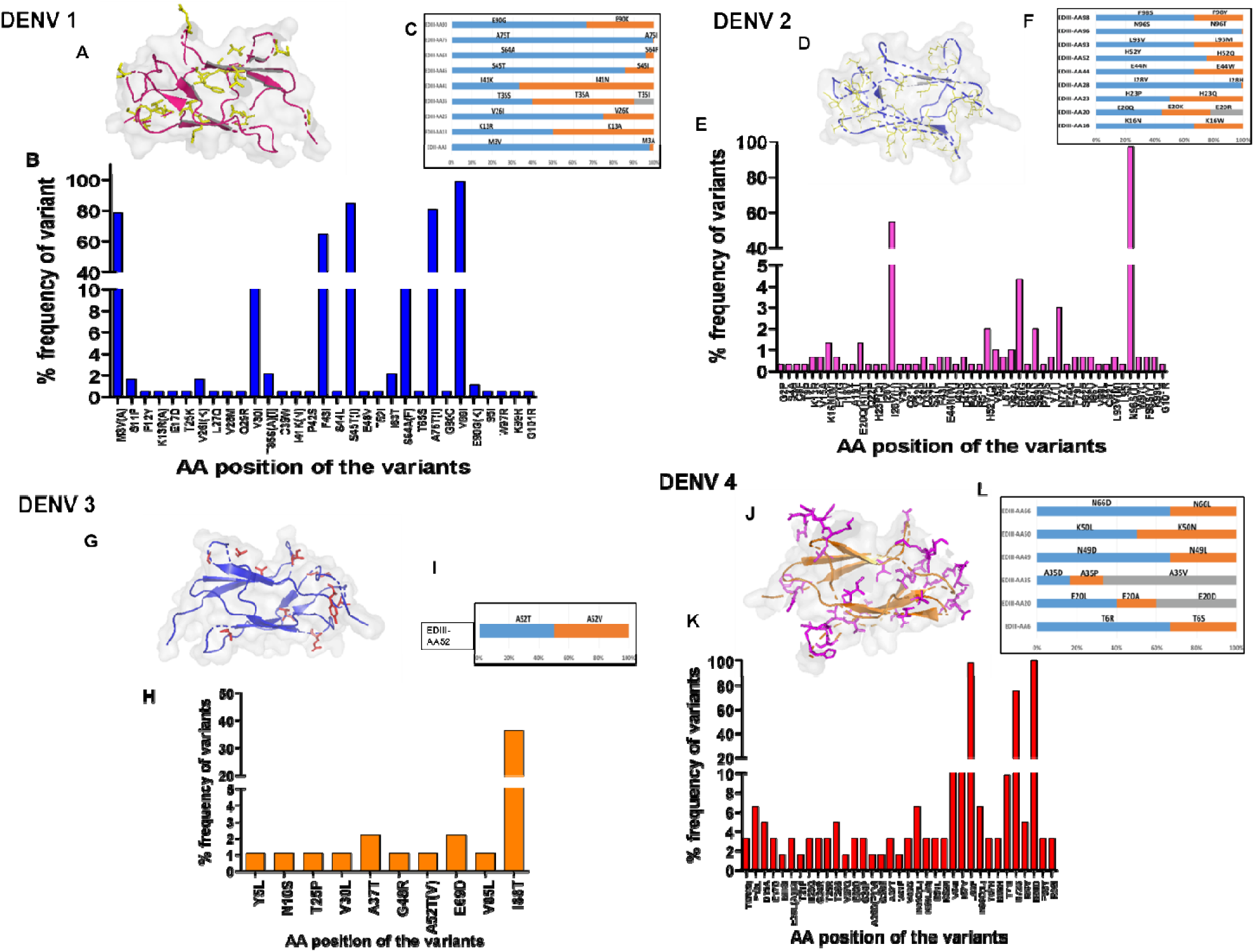
DENV amino acid variation among Indian strains of all four serotypes isolated from 2000-2020. **A, D, G & J)** The genetic diversity within EDIII of DENV1, DENV2, DENV3 and DENV4 Indian strain variants spotted on EDIII PDB structure (ribbon). Stick and ball representation on PDB structure indicates variants at the particular position. PDB: 4gt0.1, 4ut6.2.8,4Gsx.1. and 5BIC.1 used to annotate the genetic variants of DENV. **B, E, H & K)** Frequencies of variants within Indian DENV1, DENV2, DENV3, DENV4 strains, respectively. Amino acid variants residue identified relative to (NC_001477(DENV1), NC_001474 (DENV2), NC_001475 (DENV3), NC_002640 (DENV4). **C, F, I and L)** Relative frequencies of multiple amino acid substitution at a given position in DENV1, DENV2, DENV3 and DENV4 represented in different colors, respectively,

**Supplementary figure 2:**
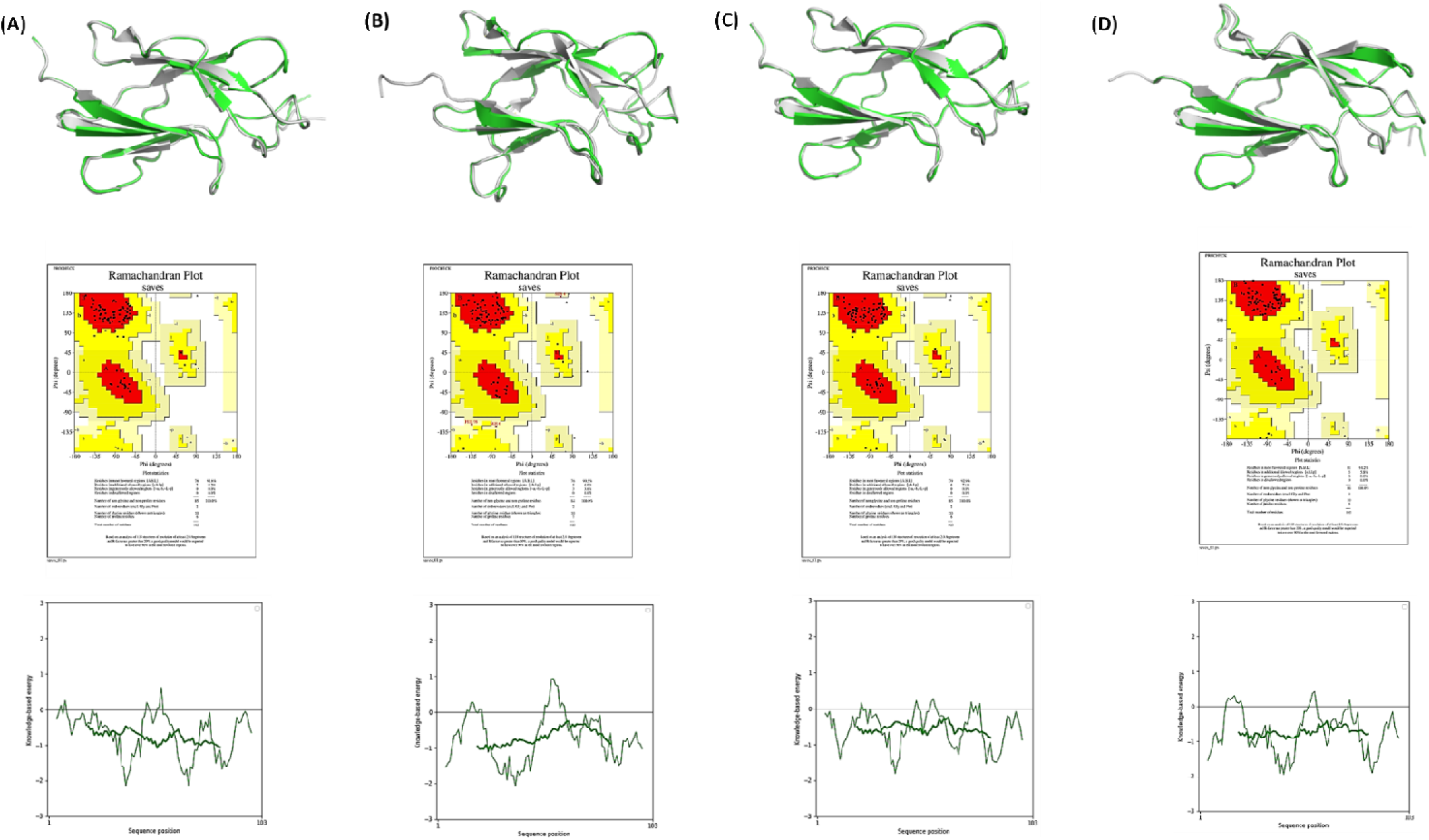
Structural validation of the modelled constructs. The modeled EDIII constructs for the serotypes (1-4) showing the structural alignment with the template, Ramachandran plot analysis and ProSA energy analysis (**A-D**). The rmsd indicates values are between 0.17-0.25, Ramachandran map shows that most of the residues are in the allowed region in the predicted structure, the predicted z-score based on ProSA analysis is negative and within the ranges for the solved crystal structures. Together, these analyses indicate that the constructs are predicted to be stable folded as the native EDIII domain in the dengue envelope protein.

**Supplementary figure 3.**
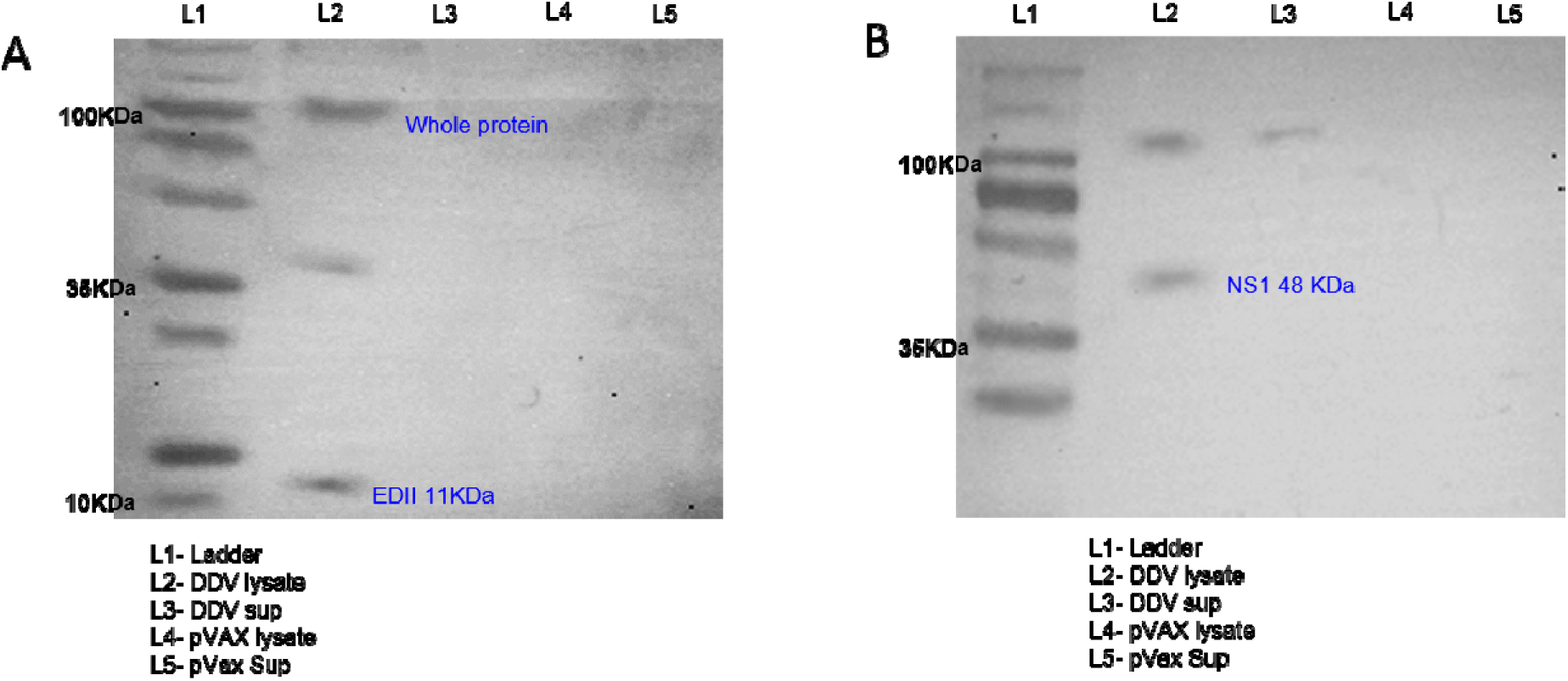
*In vitro* antigen expression using monoclonal EDIII and NS1 antibodies. Analysis of in vitro expression of ED III (**A**) and NS1 (**B**) protein after transfection of 293T cells with DDV or plasmid control by Western blot. 293T cells supernatant and lysates resolved on a gel and probed with EDIII and NS1 monoclonal antibodies. Blots were stripped then probed with β-actin loading control.

**Supplementary figure 4.**
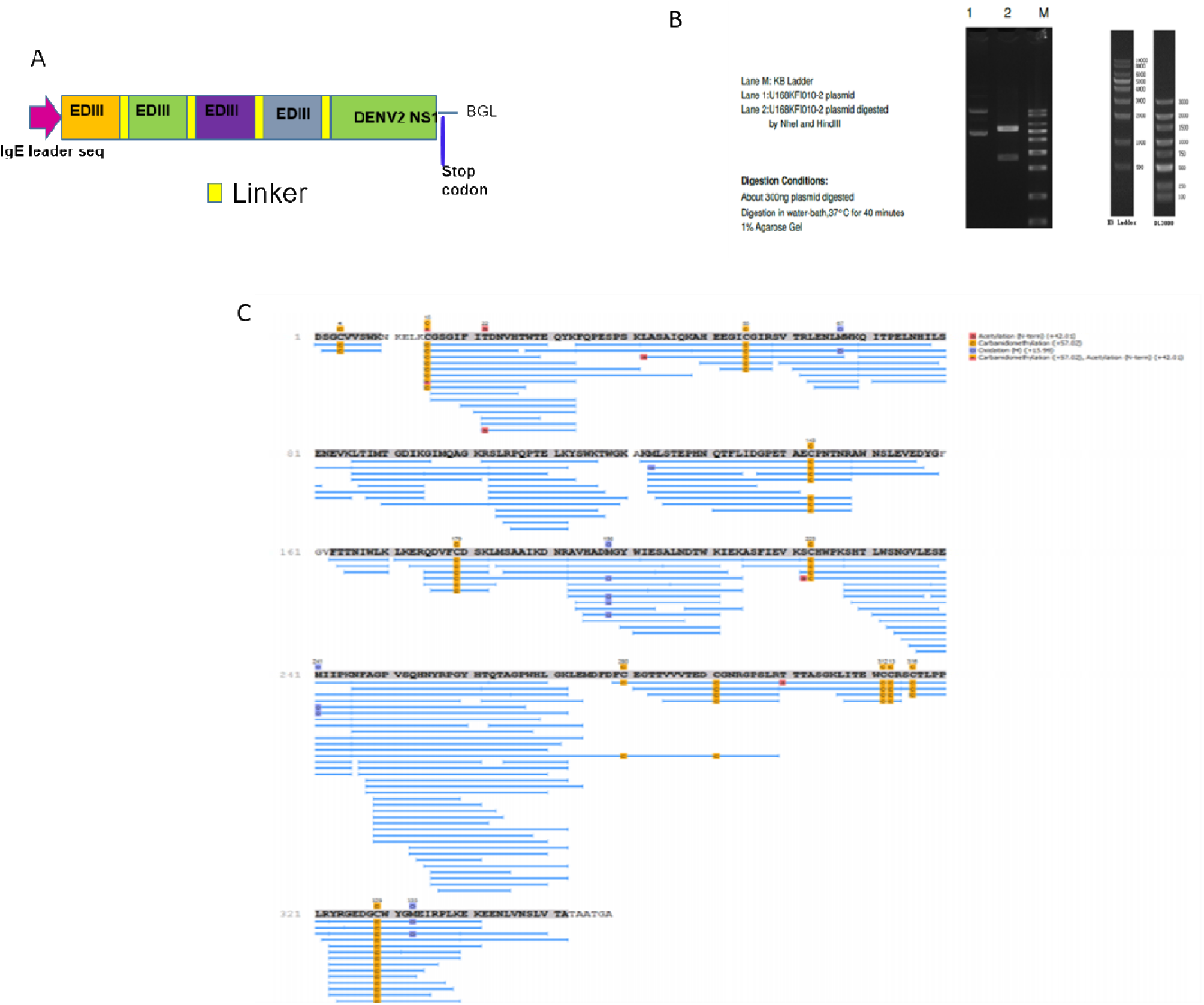
Recombinant protein synthesis. **A)** Schematic representation of recombinant protein synthesis design. **B)** Enzyme Digestion: About 300ng plasmid digested. Digestion in water-bath,37°C for 40 minutes and analysed in 1% Agarose Gel. **C)** Mass spec profile of His-purified recombinant protein

## Notes

### Competing Interest Statement

The authors have declared no competing interest.

### Author Declarations

The Institutional Ethical Clearance Review Boards approved the study of all institutions participating in this study (SJRI- IRB NO:-236/2016, AIIMS, Jodhpur-AIIMS/IEC/2017/49, & AIIMS/IEC/2019-20/939, KIH- IRB NO:07/2018, Ethics/THSTI/2011/2.1; AIIMS: IEC/NP:338/2011). The study was also approved by National Centre for Biological Sciences (NCBS) and Rajiv Gandhi Centre for Biotechnology (RGCB) Institutional Animal Ethics committee.

## References

1. Polack, F. P. et al. Safety and Efficacy of the BNT162b2 mRNA Covid-19 Vaccine. N. Engl. J. Med. 383, 2603–2615 (2020).

2. Jackson, L. A. et al. An mRNA Vaccine against SARS-CoV-2 — Preliminary Report. N. Engl. J. Med. 383, 1920–1931 (2020).

3. Silveira, M. M., Moreira, G. M. S. G. & Mendonça, M. DNA vaccines against COVID-19: Perspectives and challenges. Life Sci. 267, 118919 (2021).

4. G, B. et al. Humoral and cellular immune response against SARS-CoV-2 variants following heterologous and homologous ChAdOx1 nCoV-19/BNT162b2 vaccination. (2021) doi:10.21203/RS.3.RS-580444/V1.

5. Schmidt, T. et al. Immunogenicity and reactogenicity of heterologous ChAdOx1 nCoV-19/mRNA vaccination. Nat. Med. 2021 1–6 (2021) doi:10.1038/s41591-021-01464-w.

6. Restifo, N., Ying, H., Hwang, L. & Leitner, W. The promise of nucleic acid vaccines. Gene Ther. 7, 89 (2000).

7. Leitner, W. W., Ying, H. & Restifo, N. P. DNA and RNA-based vaccines: Principles, progress and prospects. Vaccine vol. 18 765–777 (1999).

8. Smith, T. R. F. et al. Immunogenicity of a DNA vaccine candidate for COVID-19. Nat. Commun. 11, 1–13 (2020).

9. Flingai, S. et al. Synthetic DNA vaccines: Improved vaccine potency by electroporation and co-delivered genetic adjuvants. Frontiers in Immunology vol. 4 (2013).

10. Zhang, M., Sun, J., Li, M. & Jin, X. Modified mRNA-LNP Vaccines Confer Protection against Experimental DENV-2 Infection in Mice. Mol. Ther. - Methods Clin. Dev. 18, 702–712 (2020).

11. Ramanathan, M. P. et al. Development of a novel DNA SynCon^TM^ tetravalent dengue vaccine that elicits immune responses against four serotypes. Vaccine 27, 6444–6453 (2009).

12. Pinto, P. B. A. et al. T cell responses induced by DNA vaccines based on the DENV2 e and NS1 proteins in mice: Importance in protection and immunodominant epitope identification. Front. Immunol. 10, 1522 (2019).

13. HW, C. et al. The Immunodominance Change and Protection of CD4+ T-Cell Responses Elicited by an Envelope Protein Domain III-Based Tetravalent Dengue Vaccine in Mice. PLoS One 10, e0145717–e0145717 (2015).

14. Bhatt, S. et al. The global distribution and burden of dengue. Nature 496, 504–507 (2013).

15. St. John, A. L. & Rathore, A. P. S. Adaptive immune responses to primary and secondary dengue virus infections. Nature Reviews Immunology vol. 19 218–230 (2019).

16. Murhekar, M. V., et al. Burden of dengue infection in India, 2017: a cross-sectional population based serosurvey. Lancet Glob. Heal. 7, e1065–e1073 (2019).

17. Waggoner, J. J. et al. Homotypic Dengue Virus Reinfections in Nicaraguan Children. J. Infect. Dis. 214, 986–993 (2016).

18. Forshey, B. M. et al. Incomplete Protection against Dengue Virus Type 2 Re-infection in Peru. PLoS Negl. Trop. Dis. 10, (2016).

19. Martinez, D. R. et al. Antigenic Variation of the Dengue Virus 2 Genotypes Impacts the Neutralization Activity of Human Antibodies in Vaccinees. Cell Rep. 33, 108226 (2020).

20. D, C. et al. Emergence of the Asian genotype of DENV-1 in South India. Virology 510, 40–45 (2017).

21. BP, D. et al. Circulation of different lineages of Dengue virus 2, genotype American/Asian in Brazil: dynamics and molecular and phylogenetic characterization. PLoS One 8, (2013).

22. A, M. et al. Lineage shift in Indian strains of Dengue virus serotype-3 (Genotype III), evidenced by detection of lineage IV strains in clinical cases from Kerala. Virol. J. 10, (2013).

23. Ahamed, S. F. et al. Emergence of new genotypes and lineages of dengue viruses during the 2012–15 epidemics in southern India. Int. J. Infect. Dis. 84, S34–S43 (2019).

24. M, D. et al. Complete assembly of a dengue virus type 3 genome from a recent genotype III clade by metagenomic sequencing of serum. Wellcome open Res. 3, (2019).

25. M, K. et al. Isolation and molecular characterization of dengue virus clinical isolates from pediatric patients in New Delhi. Int. J. Infect. Dis. 84S, S25–S33 (2019).

26. Alagarasu, K. et al. Serotype and genotype diversity of dengue viruses circulating in India: a multi-centre retrospective study involving the Virus Research Diagnostic Laboratory Network in 2018. Int. J. Infect. Dis. 111, 242–252 (2021).

27. Shrivastava, S. et al. Co-circulation of all the four dengue virus serotypes and detection of a novel clade of DENV-4 (genotype I) virus in Pune, India during 2016 season. PLoS One 13, e0192672 (2018).

28. Masika, M. M. et al. Detection of dengue virus type 2 of Indian origin in acute febrile patients in rural Kenya. PLoS Negl. Trop. Dis. 14, e0008099 (2020).

29. Gathii, K., Nyataya, J. N., Mutai, B. K., Awinda, G. & Waitumbi, J. N. Complete Coding Sequences of Dengue Virus Type 2 Strains from Febrile Patients Seen in Malindi District Hospital, Kenya, during the 2017 Dengue Fever Outbreak. Genome Announc. 6, (2018).

30. Dengue vaccine: WHO position paper, September 2018 – Recommendations. Vaccine vol. 37 4848–4849 (2019).

31. Juraska, M. et al. Viral genetic diversity and protective efficacy of a tetravalent dengue vaccine in two phase 3 trials. Proc. Natl. Acad. Sci. U. S. A. 115, E8378–E8387 (2018).

32. Holmes, E. C. & Twiddy, S. S. The origin, emergence and evolutionary genetics of dengue virus. Infection, Genetics and Evolution vol. 3 19–28 (2003).

33. Rico-Hesse, R. Molecular evolution and distribution of dengue viruses type 1 and 2 in nature. Virology 174, 479–493 (1990).

34. Frei, J. C. et al. Engineered Dengue Virus Domain III Proteins Elicit Cross-Neutralizing Antibody Responses in Mice. J. Virol. 92, (2018).

35. Li, X. Q. et al. Dengue virus envelope domain III immunization elicits predominantly cross-reactive, poorly neutralizing antibodies localized to the AB loop: Implications for dengue vaccine design. J. Gen. Virol. 94, 2191–2201 (2013).

36. Ramasamy, V. et al. A tetravalent virus-like particle vaccine designed to display domain III of dengue envelope proteins induces multi-serotype neutralizing antibodies in mice and macaques which confer protection against antibody dependent enhancement in AG129 mice. PLoS Negl. Trop. Dis. 12, (2018).

37. Rajpoot, R. K., Shukla, R., Arora, U., Swaminathan, S. & Khanna, N. Dengue envelope-based ‘four-in-one’ virus-like particles produced using Pichia pastoris induce enhancement-lacking, domain III-directed tetravalent neutralising antibodies in mice. Sci. Rep. 8, (2018).

38. Islam, M. M., Miura, S., Hasan, M. N., Rahman, N. & Kuroda, Y. Anti-Dengue ED3 Long-Term Immune Response With T-Cell Memory Generated Using Solubility Controlling Peptide Tags. Front. Immunol. 0, 333 (2020).

39. Gallichotte, E. N. et al. Role of Zika Virus Envelope Protein Domain III as a Target of Human Neutralizing Antibodies. (2019) doi:10.1128/mBio.01485-19.

40. Chávez, J. H., Silva, J. R., Amarilla, A. A. & Moraes Figueiredo, L. T. Domain III peptides from flavivirus envelope protein are useful antigens for serologic diagnosis and targets for immunization. Biologicals vol. 38 613–618 (2010).

41. Robinson, L. N. et al. Structure-Guided Design of an Anti-dengue Antibody Directed to a Non-immunodominant Epitope. Cell 162, 493–504 (2015).

42. Gromowski, G. D., Barrett, N. D. & Barrett, A. D. T. Characterization of Dengue Virus Complex-Specific Neutralizing Epitopes on Envelope Protein Domain III of Dengue 2 Virus. J. Virol. 82, 8828–8837 (2008).

43. GD, G. et al. Mutations of an antibody binding energy hot spot on domain III of the dengue 2 envelope glycoprotein exploited for neutralization escape. Virology 407, 237–246 (2010).

44. CW, L. & SC, W. A functional epitope determinant on domain III of the Japanese encephalitis virus envelope protein interacted with neutralizing-antibody combining sites. J. Virol. 77, 2600–2606 (2003).

45. X, Z. et al. Structures and Functions of the Envelope Glycoprotein in Flavivirus Infections. Viruses 9, (2017).

46. Weaver, E. A., Rubrum, A. M., Webby, R. J. & Barry, M. A. Protection against Divergent Influenza H1N1 Virus by a Centralized Influenza Hemagglutinin. PLoS One 6, e18314 (2011).

47. Meyerhoff, R. R. et al. HIV-1 Consensus Envelope-Induced Broadly Binding Antibodies. AIDS Res. Hum. Retroviruses 33, 859–868 (2017).

48. Wang, R. et al. Vaccination With a Single Consensus Envelope Protein Ectodomain Sequence Administered in a Heterologous Regimen Induces Tetravalent Immune Responses and Protection Against Dengue Viruses in Mice. Front. Microbiol. 0, 1113 (2019).

49. M, M. et al. A consensus epitope prediction approach identifies the breadth of murine T(CD8+)-cell responses to vaccinia virus. Nat. Biotechnol. 24, 817–819 (2006).

50. Yan, J. et al. Enhanced cellular immune responses elicited by an engineered HIV-1 subtype B consensus-based envelope DNA vaccine. Mol. Ther. 15, 411–421 (2007).

51. Chen, H. R. et al. Dengue Virus Nonstructural Protein 1 Induces Vascular Leakage through Macrophage Migration Inhibitory Factor and Autophagy. PLoS Negl. Trop. Dis. 10, (2016).

52. Chen, H. R., Lai, Y. C. & Yeh, T. M. Dengue virus non-structural protein 1: A pathogenic factor, therapeutic target, and vaccine candidate. Journal of Biomedical Science vol. 25 (2018).

53. Wan, S. W. et al. Protection against dengue virus infection in mice by administration of antibodies against modified nonstructural protein 1. PLoS One 9, (2014).

54. Beatty, P. R. et al. Dengue virus NS1 triggers endothelial permeability and vascular leak that is prevented by NS1 vaccination. Sci. Transl. Med. 7, 304ra141–304ra141 (2015).

55. Modis, Y. Relating structure to evolution in class II viral membrane fusion proteins. Current Opinion in Virology vol. 5 34–41 (2014).

56. Malavige, G. N. et al. Identification of serotype-specific T cell responses to highly conserved regions of the dengue viruses. Clin. Exp. Immunol. 168, 215 (2012).

57. JV, K., C, L., O, L. & M, N. Reliable B cell epitope predictions: impacts of method development and improved benchmarking. PLoS Comput. Biol. 8, (2012).

58. B, R., B, A., S, P., B, P. & M, N. NetMHCpan-4.1 and NetMHCIIpan-4.0: improved predictions of MHC antigen presentation by concurrent motif deconvolution and integration of MS MHC eluted ligand data. Nucleic Acids Res. 48, W449–W454 (2020).

59. Wang, P. et al. Peptide binding predictions for HLA DR, DP and DQ molecules. BMC Bioinforma. 2010 111 11, 1–12 (2010).

60. N, M., Y, A. & K, T. Foreign gene expression in an organotypic culture of cortical anlage after in vivo electroporation. Neuroreport 10, 2319–2323 (1999).

61. Itasaki, N., Bel-Vialar, S. & Krumlauf, R. ‘Shocking’ developments in chick embryology: Electroporation and in ovo gene expression. Nat. Cell Biol. 1, (1999).

62. ML, B. et al. Immunotherapy against HPV16/18 generates potent TH1 and cytotoxic cellular immune responses. Sci. Transl. Med. 4, (2012).

63. A, F., J, D. C. & D, Z. A statistically defined endpoint titer determination method for immunoassays. J. Immunol. Methods 221, 35–41 (1998).

64. S, S., MG, A. & E, S. Comparative whole genome analysis of dengue virus serotype-2 strains differing in trans-endothelial cell leakage induction in vitro. Infect. Genet. Evol. 52, 34–43 (2017).

65. Singla, M. et al. Immune Response to Dengue Virus Infection in Pediatric Patients in New Delhi, India—Association of Viremia, Inflammatory Mediators and Monocytes with Disease Severity. PLoS Negl. Trop. Dis. 10, e0004497 (2016).

66. Shastri, J., Williamson, M., Vaidya, N., Agrawal, S. & Shrivastav, O. Nine year trends of dengue virus infection in Mumbai, Western India. J. Lab. Physicians 9, 296–302 (2017).

67. DY, C. et al. Strategically examining the full-genome of dengue virus type 3 in clinical isolates reveals its mutation spectra. Virol. J. 2, (2005).

68. GN, R., C, R. & V, S. Stereochemistry of polypeptide chain configurations. J. Mol. Biol. 7, 95–99 (1963).

69. Wiederstein, M. & Sippl, M. J. ProSA-web: interactive web service for the recognition of errors in three-dimensional structures of proteins. Nucleic Acids Res. 35, W407–W410 (2007).

70. JT, R., J, H. & AD, B. Guidelines for Plaque-Reduction Neutralization Testing of Human Antibodies to Dengue Viruses. Viral Immunol. 21, 123–132 (2008).

71. Feng, K. et al. Long-Term Protection Elicited by a DNA Vaccine Candidate Expressing the prM-E Antigen of Dengue Virus Serotype 3 in Mice. Front. Cell. Infect. Microbiol. 10, 87 (2020).

72. Becerra, A. et al. Increased activity of indoleamine 2,3-dioxygenase in serum from acutely infected dengue patients linked to gamma interferon antiviral function. J. Gen. Virol. 90, 810 (2009).

73. Diamond, M. S. et al. Modulation of Dengue Virus Infection in Human Cells by Alpha, Beta, and Gamma Interferons. J. Virol. 74, 4957 (2000).

74. Rosenthal, K. S. & Zimmerman, D. H. Vaccines: All things considered. Clinical and Vaccine Immunology vol. 13 821–829 (2006).

75. Smatti, M. K., Al Thani, A. A. & Yassine, H. M. Viral-induced enhanced disease illness. Front. Microbiol. 9, 2991 (2018).

76. R, de A. et al. A single dose of self-transcribing and replicating RNA-based SARS-CoV-2 vaccine produces protective adaptive immunity in mice. Mol. Ther. 29, 1970–1983 (2021).

77. Krathwohl, M. D. & Anderson, J. L. Chemokine CXCL10 (IP-10) is sufficient to trigger an immune response to injected antigens in a mouse model. Vaccine 24, 2987–2993 (2006).

78. Inoue, H. & Kubo, M. SOCS proteins in T helper cell differentiation: Implications for allergic disorders? Expert Reviews in Molecular Medicine vol. 6 (2004).

79. Yoshimura, A., Suzuki, M., Sakaguchi, R., Hanada, T. & Yasukawa, H. SOCS, inflammation, and autoimmunity. Frontiers in Immunology vol. 3 (2012).

80. Ford, J. et al. CCL7 is a negative regulator of cutaneous inflammation following Leishmania major infection. Front. Immunol. 10, (2019).

81. Mahmood, N., Mihalcioiu, C. & Rabbani, S. A. Multifaceted role of the urokinase-type plasminogen activator (uPA) and its receptor (uPAR): Diagnostic, prognostic, and therapeutic applications. Frontiers in Oncology vol. 8 (2018).

82. Andresen, E., Günther, G., Bullwinkel, J., Lange, C. & Heine, H. Increased expression of beta-defensin 1 (DEFB1) in chronic obstructive pulmonary disease. PLoS One 6, (2011).

83. Loyet, K. M., Ouyang, W., Eaton, D. L. & Stults, J. T. Proteomic profiling of surface proteins on Th1 and Th2 Cells. J. Proteome Res. 4, 400–409 (2005).

84. Urata, S. et al. BST-2 controls T cell proliferation and exhaustion by shaping the early distribution of a persistent viral infection. PLoS Pathog. 14, (2018).

85. Salem, M., Mony, J. T., Løbner, M., Khorooshi, R. & Owens, T. Interferon regulatory factor-7 modulates experimental autoimmune encephalomyelitis in mice. J. Neuroinflammation 8, (2011).

86. Pollett, S. et al. Understanding dengue virus evolution to support epidemic surveillance and counter-measure development. Infect. Genet. Evol. 62, 279–295 (2018).

87. Katzelnick, L. C. et al. Antibody-dependent enhancement of severe dengue disease in humans. Science (80-. ). 358, 929–932 (2017).

88. Martinez, D. R. et al. Antigenic Variation of the Dengue Virus 2 Genotypes Impacts the Neutralization Activity of Human Antibodies in Vaccinees. Cell Rep. 33, 108226 (2020).

89. Sukupolvi-Petty, S. et al. Structure and Function Analysis of Therapeutic Monoclonal Antibodies against Dengue Virus Type 2. J. Virol. 84, 9227–9239 (2010).

90. B, S. et al. The development of therapeutic antibodies that neutralize homologous and heterologous genotypes of dengue virus type 1. PLoS Pathog. 6, 1–18 (2010).

91. Brien, J. D. et al. Genotype-Specific Neutralization and Protection by Antibodies against Dengue Virus Type 3. J. Virol. 84, 10630 (2010).

92. Wahala, W. M. P. B. et al. Natural strain variation and antibody neutralization of dengue serotype 3 viruses. PLoS Pathog. 6, 1000821 (2010).

93. Zhou, Y. et al. The mechanism of differential neutralization of dengue serotype 3 strains by monoclonal antibody 8A1. Virology 439, 57 (2013).

94. Wahala, W. M. P. B. et al. Natural strain variation and antibody neutralization of dengue serotype 3 viruses. PLoS Pathog. 6, 1000821 (2010).

95. Hobernik, D. & Bros, M. DNA Vaccines—How Far From Clinical Use? Int. J. Mol. Sci. 19, (2018).

96. Muthumani, K. et al. Immunogenicity of novel consensus-based DNA vaccines against Chikungunya virus. Vaccine 26, 5128–5134 (2008).

97. Tebas, P. et al. Safety and Immunogenicity of an Anti–Zika Virus DNA Vaccine — Preliminary Report. N. Engl. J. Med. (2017) doi:10.1056/nejmoa1708120.

98. Modjarrad, K. et al. Safety and immunogenicity of an anti-Middle East respiratory syndrome coronavirus DNA vaccine: a phase 1, open-label, single-arm, dose-escalation trial. Lancet Infect. Dis. 19, 1013–1022 (2019).

99. Tebas, P., et al. Safety and immunogenicity of INO-4800 DNA vaccine against SARS-CoV-2: A preliminary report of an open-label, Phase 1 clinical trial. EClinicalMedicine 31, (2021).

100. Dey, A. et al. Immunogenic potential of DNA vaccine candidate, ZyCoV-D against SARS-CoV-2 in animal models. Vaccine 39, 4108 (2021).

101. Filipe, A. R., Martins, C. M. V & Rociia, H. Laboratory Infection with Zika Virus after Vaccination against Yellow Fever.

102. Wieten, R. W. et al. A Single 17D Yellow Fever Vaccination Provides Lifelong Immunity; Characterization of Yellow-Fever-Specific Neutralizing Antibody and T-Cell Responses after Vaccination. PLoS One 11, e0149871 (2016).

103. Maciejewski, S. et al. Distinct neutralizing antibody correlates of protection among related Zika virus vaccines identify a role for antibody quality. Sci. Transl. Med. 12, 9066 (2020).

104. Weiskopf, D. et al. The Human CD8 + T Cell Responses Induced by a Live Attenuated Tetravalent Dengue Vaccine Are Directed against Highly Conserved Epitopes . J. Virol. 89, 120–128 (2015).

105. Hellerstein, M. What are the roles of antibodies versus a durable, high quality T-cell response in protective immunity against SARS-CoV-2? Vaccine X 6, 100076 (2020).

106. Cox, M. A., Kahan, S. M. & Zajac, A. J. Anti-viral CD8 T cells and the cytokines that they love. Virology vol. 435 157–169 (2013).

107. Huber, J. P. & David Farrar, J. Regulation of effector and memory T-cell functions by type I interferon. Immunology vol. 132 466–474 (2011).

108. McNab, F., Mayer-Barber, K., Sher, A., Wack, A. & O’Garra, A. Type I interferons in infectious disease. Nature Reviews Immunology vol. 15 87–103 (2015).

109. Costa, V. V., Fagundes, C. T., Souza, D. G. & Teixeira, M. M. Inflammatory and Innate Immune Responses in Dengue Infection: Protection versus Disease Induction. Am. J. Pathol. 182, 1950–1961 (2013).

110. Foulds, K. E., Wu, C. Y. & Seder, R. A. Th1 memory: Implications for vaccine development. Immunological Reviews vol. 211 58–66 (2006).

111. Lukacs, N. W. & Malinczak, C. A. Harnessing cellular immunity for vaccination against respiratory viruses. Vaccines vol. 8 1–21 (2020).

