## Supplementary Table 1-4 for "Immune profile and responses of a novel Dengue DNA vaccine encoding EDIII-NS1 consensus design based on Indo-African sequences"

**Supplementary Table 1: Clinical characteristics and biochemical correlates of sequenced samples**

| **Variables / Characteristics*** | **All patients (N=319)^a^** | **Dengue Fever (N=114)** | **Dengue with warning signs N=128** | **Severe dengue N=28** | **p value** | **Statistical test** |
| --- | --- | --- | --- | --- | --- | --- |
| **Patient characteristics** | | | | | | |
| Age | 23 (0.6, 83) | 24 (4,66) | 25 (0.66,83) | 21 (0.75, 69) | 0.09 | Kruskal Wallis test |
| **Sex** | | | | | | |
| Female; n (%) | 140 (43.8) | 43 (37.7) | 58 (45.3) | 9 (32.1) | 0.42 | Chi-square test |
| Male; n (%) | 173 (54.23) | 57 (50) | 57 (44.5) | 14 (50) |  |  |
| **Onset symptoms** | | | | | | |
| Average days of fever; mean ±SD | 3.35±1.96 | 2.89±1.38 | 3.41±1.5 | 3.42±1.34 | 0.35 | ANOVA test |
| Abdominal Pain; n (%) | 54 (16.92) | 13 (20.63) | 15 (13.27) | 10 (35.71) | **0.021** | **Chi-square test** |
| Rash; n (%) | 26 (8.15) | 10 (17.24) | 12 (19.04) | 4 (21.05) | 0.92 | Chi-square test |
| Retro Orbital Pain; n (%) | 54 (16.92) | 22 (35.55) | 16 (34.04) | 8 (47.05) | 0.431 | Chi-square test |
| Hepatomegaly; n (%) | 30 (9.4) | 11 (24.13) | 8 (20.68) | 3 (22.22) | 0.77 | Chi-square test |
| **Serological classification** | | | | | | |
| Primary infection; n(%) | 191 (59.8) | 72 (63.1) | 70 (54.68) | 13 (46.42) | **0.024** | **Chi-square test** |
| Secondary infection; n(%) | 62 (19.43) | 17 (14.91) | 29 (22.65) | 11 (39.28) |  |  |
| Unknown; n(%)**^b^** | 66 (20.68) | 25(21.92) | 29(22.65) | 4(14.28) |  |  |
| **Biochemical parameters** | | | | | | |
| Hemoglobin (g/dL) mean (SD), | 12.79±2.09 | 12.85±2.21 | 12.89±1.91 | 11.87±2.35 | 0.06 | ANOVA test |
| TC /cumm | 4300 (1260,18800) | 4600 (2000,15400) N=101 | 4100 (1260,18800) N=121 | 4415 (2000, 10010) N=28 | 0.2657 | Kruskal Wallis |
| Hematocrit; %. mean (SD) | 37.70 ± 5.51 | 35.97 ±4.90  N=55 | 38.0 ± 4.62 N=55 | 42.48 ± 7.38 N=16 | **<0.0001** | ANOVA test |
| Platelet count at admission/ cumm |  | 136 (41,302) N=105 | 97 (11,346) N=115 | 39 (14,225) N=28 | **4.398E-11 (<0.0001)** | Kruskall Wallis |
| Platelet count (least)/cumm |  | 82 (38,185) N=13 | 31 (6, 182) N=41 | 32 (15, 45) N=7 | **0.000814** | Kruskall Wallis |
| Aspartate aminotransferase; IU/L | 52.5 (10, 1480) N=206 | 38 (10, 673) N=73 | 66 (13, 562) N=105 | 150 (30, 1480) N=24 | **0.0000000003102  (<0.0001)** | Kruskall Wallis |
| Alanine aminotransferase; IU/L | 40 (12,802) N=206 | 32 (13, 743) N=73 | 41 (12, 454) N=105 | 87 (13, 802) N=24 | **0.0001922** | Kruskall Wallis |

*****Values are Median unless stated otherwise. SD- standard deviation

**^a^** Total number of Samples sequenced: Since the lack of clinical and Biochemical correlates WHO classification was not done for 49 cases

**^b^** Unknown baseline serostatus across stratifications: Inadequate samples to perform serum dengue-specific IgM and IgG Elisa

**Supplementary Table 2**

Table 1

EDIII diversity in DENV 1-4 genotype variants

| DENV1 | | DENV2 | | DENV3 | | DENV4 | |
| --- | --- | --- | --- | --- | --- | --- | --- |
| Genotype 1 | 4.85 (2.91-10.68) | Cosmopolitan | 1.94        (4.85 -6.8) | Genotype 1 | 5.83 (4.85-6.8) | Genotype 1 | 3.88  (1.94-9.71) |
| Genotype 2 | 1.94 (1.94-2.91) | American | 1.94** (0.97-2.91) | Genotype 2 | 2.91 (2.91-4.85) | Genotype 2 | 2.91  (2.91-3.88) |
| Genotype 3 | 3.88 (1.94-4.85) | Asian 1 | 1.94  (0.97-1.94) | Genotype 3 | 1.94 (0.97-3.88) | Genotype 3 | 3.88 *** (3.88-4.85) |
| Genotype 4 | 3.88 | Asian 2 | 0 (0-0.97) | Genotype 4 | 4.85  (4.85-6.8) | Genotype 5 | 3.39 (1.94-11.65) |
| Genotype 5 | 4.85  (2.91-6.8) | Asian American | 0.97 (0.97-2.91) |  | | Sylvatic | 8.74 |
| Sylvatic | 1.94 | Sylvatic | 4.85 (2.91-6.8) |  |  |  | |
| **1 seq had 100% identity    ***7 seq had 100% identity | | | | | | | |

**Supplementary Table 3**

**Comparison of multiple EDIIIs within each serotype with the EDIII consensus sequences used to design DENV DNA vaccine**

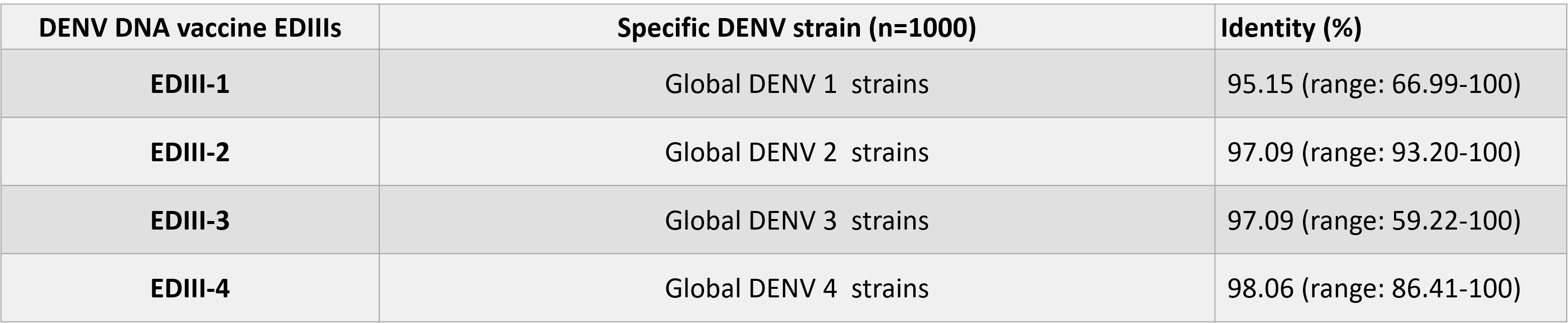

**DENV EDIII sequences used in designing DDV were aligned with the global dengue sequences for each serotype (n=1000) in the VIPR database to obtain the range of percent identity within members of the given serotype.**

**DENV EDIII sequences used in designing DDV were aligned with the global dengue sequences for each serotype (n=1000) in the VIPR database to obtain the range of percent identity within members of the given serotype.**

**Supplementary Table 4a-d: B-cell structural epitopes predicted for the constructs across serotypes.**

DiscoTope tool implemented in the IEDB website was used for the analysis.

**Supplementary Table 4a: Predictions for the Serotype 1 construct sequence**

| **Residue ID** | **Residue Name** | **Contact Number** | **Propensity Score** | **Discotope Score** |
| --- | --- | --- | --- | --- |
| 317 | HIS | 14 | -0.622 | -7.622 |
| 342 | GLU | 16 | 0.822 | -7.178 |
| 343 | LYS | 10 | 1.189 | -3.811 |
| 344 | GLY | 11 | 1.67 | -3.83 |
| 345 | VAL | 13 | 1.881 | -4.619 |
| 346 | THR | 13 | 1.482 | -5.018 |
| 347 | GLN | 14 | 2.237 | -4.763 |
| 348 | ASN | 11 | 1.448 | -4.052 |
| 349 | GLY | 14 | 1.247 | -5.753 |
| 362 | GLU | 12 | 1.659 | -4.341 |
| 363 | LYS | 13 | 0.749 | -5.751 |
| 374 | GLY | 13 | -0.101 | -6.601 |
| 394 | LYS | 12 | 0.257 | -5.743 |
| 395 | GLY | 10 | 0.482 | -4.518 |
| 396 | SER | 11 | -0.217 | -5.717 |
| 397 | SER | 11 | -0.313 | -5.813 |

**Supplementary Table 4b: Predictions for the Serotype 2 construct sequence**

| **Chain ID** | **Residue ID** | **Residue Name** | **Contact Number** | **Propensity Score** | **Discotope Score** |
| --- | --- | --- | --- | --- | --- |
| A | 311 | GLU | 11 | -1.429 | -6.929 |
| A | 318 | GLY | 16 | 0.341 | -7.659 |
| A | 327 | GLU | 12 | -0.658 | -6.658 |
| A | 329 | ASP | 13 | -1.091 | -7.591 |
| A | 342 | LEU | 13 | -0.803 | -7.303 |
| A | 343 | GLU | 9 | -0.548 | -5.048 |
| A | 345 | ARG | 9 | -0.09 | -4.59 |
| A | 346 | HIS | 12 | -0.739 | -6.739 |
| A | 360 | GLU | 15 | 1.117 | -6.383 |
| A | 362 | ASP | 12 | 1.48 | -4.52 |
| A | 363 | SER | 16 | 1.569 | -6.431 |
| A | 373 | PHE | 14 | 0.264 | -6.736 |
| A | 374 | GLY | 11 | 0.583 | -4.917 |
| A | 375 | ASP | 15 | 0.462 | -7.038 |
| A | 383 | GLU | 11 | -2.124 | -7.624 |
| A | 384 | PRO | 10 | -1.814 | -6.814 |

**Supplementary Table 4c: Predictions for the Serotype 3 construct sequence**

| **Chain ID** | **Residue ID** | **Residue Name** | **Contact Number** | **Propensity Score** | **Discotope Score** |
| --- | --- | --- | --- | --- | --- |
| A | 317 | HIS | 14 | -0.622 | -7.622 |
| A | 342 | GLU | 16 | 0.822 | -7.178 |
| A | 343 | LYS | 10 | 1.189 | -3.811 |
| A | 344 | GLY | 11 | 1.67 | -3.83 |
| A | 345 | VAL | 13 | 1.881 | -4.619 |
| A | 346 | THR | 14 | 0.973 | -6.027 |
| A | 347 | GLN | 14 | 2.237 | -4.763 |
| A | 348 | ASN | 11 | 1.448 | -4.052 |
| A | 349 | GLY | 14 | 1.247 | -5.753 |
| A | 362 | GLU | 12 | 1.659 | -4.341 |
| A | 363 | LYS | 13 | 0.749 | -5.751 |
| A | 374 | GLY | 13 | -0.101 | -6.601 |

**Supplementary Table 4d: Predictions for the Serotype 4 construct sequence**

| **Residue ID** | **Residue Name** | **Contact Number** | **Propensity Score** | **Discotope Score** |
| --- | --- | --- | --- | --- |
| 313 | THR | 12 | 0.558 | -5.442 |
| 314 | GLN | 8 | 0.512 | -3.488 |
| 315 | HIS | 8 | 0.238 | -3.762 |
| 316 | GLY | 11 | 0.538 | -4.962 |
| 340 | VAL | 16 | 1.087 | -6.913 |
| 341 | ASN | 10 | 0.871 | -4.129 |
| 342 | LYS | 13 | 0.693 | -5.807 |
| 343 | GLU | 13 | 1.073 | -5.427 |
| 344 | LYS | 15 | 0.508 | -6.992 |
| 345 | VAL | 17 | -0.678 | -9.178 |
| 346 | VAL | 11 | -0.114 | -5.614 |
| 347 | GLY | 10 | -0.547 | -5.547 |
| 352 | SER | 9 | -1.881 | -6.381 |
| 371 | PHE | 14 | 1.85 | -5.15 |
| 372 | GLY | 13 | 1.766 | -4.734 |
| 373 | ASP | 17 | 1.623 | -6.877 |
| 382 | ASN | 8 | -3.355 | -7.355 |

**Supplementary Table 5a-d: Predicted T-cell epitopes (MHCI) and the corresponding HLA alleles**

We predicted the MHCI epitopes from the four serotype EDIII construct sequence using the NetMHCpan (v4.0) implemented in IEDB.

**Supplementary Table 5a: Predictions for the Serotype 1 construct sequence**

| **MHCI epitope** | **HLA allele** |
| --- | --- |
| AETQHGTVL | HLA-B*40:01, HLA-B*44:02, HLA-B*44:03 |
| AETQHGTVLV | HLA-B*40:01 |
| AGEKALKLSW | HLA-B*44:02, HLA-B*44:03, HLA-B*57:01 |
| ALKLSWFKK | HLA-A*03:01, HLA-A*30:01 |
| CTGSFKLEK | HLA-A*11:01 |
| EPPFGESYI | HLA-B*51:01 |
| ETEPPFGESY | HLA-A*01:01, HLA-A*26:01, HLA-B*44:02, HLA-B*44:03 |
| ETQHGTVLV | HLA-A*68:02 |
| EVAETQHGT | HLA-A*68:02 |
| EVAETQHGTV | HLA-A*68:02 |
| GEKALKLSW | HLA-B*44:02, HLA-B*44:03 |
| GEKALKLSWF | HLA-B*44:02, HLA-B*44:03 |
| GTVLVQVKY | HLA-A*01:01, HLA-A*26:01, HLA-A*30:02, HLA-A*32:01, HLA-B*15:01, HLA-B*58:01 |
| HGTVLVQVKY | HLA-A*26:01 |
| ITANPIVTDK | HLA-A*03:01, HLA-A*11:01, HLA-A*30:01, HLA-A*68:01 |
| KALKLSWFK | HLA-A*03:01, HLA-A*11:01, HLA-A*30:01, HLA-A*31:01 |
| RLITANPIV | HLA-A*02:01, HLA-A*02:03, HLA-A*02:06 |
| SYVMCTGSF | HLA-A*23:01, HLA-A*24:02 |
| TANPIVTDK | HLA-A*11:01, HLA-A*68:01 |
| TEPPFGESY | HLA-B*44:02, HLA-B*44:03 |
| TQHGTVLVQV | HLA-A*02:06 |
| TQNGRLITA | HLA-A*02:03, HLA-A*02:06 |
| VAETQHGTVL | HLA-B*40:01 |
| YVMCTGSF | HLA-A*11:01 |

**Supplementary Table 5b: Predictions for the Serotype 2 construct sequence**

| **MHCI epitope** | **HLA allele** |
| --- | --- |
| AEPPFGDSY | HLA-B*44:02, HLA-B*44:03 |
| AETQHGTIV | HLA-B*40:01, HLA-B*44:02, HLA-B*44:03 |
| AETQHGTIVV | HLA-B*40:01 |
| DLEKRHVLGR | HLA-A*33:01 |
| EAEPPFGDSY | HLA-A*01:01, HLA-A*26:01, HLA-B*35:0, HLA-B*44:02, HLA-B*44:03 |
| EIAETQHGT | HLA-A*68:02 |
| EIAETQHGTI | HLA-A*68:02 |
| EIMDLEKRHV | HLA-A*68:02 |
| EPGQLKLSW | HLA-B*53:01 |
| EPGQLKLSWF | HLA-B*53:01 |
| ETQHGTIVV | HLA-A*68:02 |
| ETQHGTIVVR | HLA-A*33:01, HLA-A*68:01 |
| GQLKLSWFK | HLA-A*03:01, HLA-A*11:01 |
| GTIVVRVQY | HLA-A*11:01, HLA-A*26:0, HLA-A*30:0, HLA-A*32:0, HLA-B*15:0, HLA-B*57:0, HLA-B*58:01 |
| HGTIVVRVQY | HLA-A*26:01 |
| HVLGRLITV | HLA-A*02:01, HLA-A*02:03, HLA-A*02:06, HLA-A*32:01, HLA-A*68:01, HLA-B*08:01 |
| IIGVEPGQLK | HLA-A*03:01, HLA-A*11:01 |
| IMDLEKRHVL | HLA-B*08:01 |
| ITVNPIVTEK | HLA-A*03:01, HLA-A*11:01, HLA-A*30:01, HLA-A*68:01 |
| MDLEKRHVL | HLA-B*08:01 |
| MSYSMCTGK | HLA-A*03:01, HLA-A*11:01 |
| QLKLSWFKK | HLA-A*03:01, HLA-A*30:01 |
| RHVLGRLITV | HLA-A*02:06 |
| RLITVNPIV | HLA-A*02:01, HLA-A*02:03, HLA-A*02:06 |
| SPCKIPFEI | HLA-B*07:02, HLA-B*51:01, HLA-B*53:01 |
| SPCKIPFEIM | HLA-B*07:02 |
| SYSMCTGKF | HLA-A*23:01, HLA-A*24:02 |
| TEKDSPVNI | HLA-B*40:01, HLA-B*44:02, HLA-B*44:03 |
| TQHGTIVVR | HLA-A*31:01 |
| TVNPIVTEK | HLA-A*03:01, HLA-A*11:01, HLA-A*30:01, HLA-A*31:01, HLA-A*33:01, HLA-A*68:01 |
| VEPGQLKLSW | HLA-B*44:02, HLA-B*44:03, HLA-B*53:01 |
| YEGDGSPCKI | HLA-B*40:01 |

**Supplementary Table 5c: Predictions for the Serotype 3 construct sequence**

| **MHCI epitope** | **HLA allele** |
| --- | --- |
| AEPPFGESNI | HLA-B*40:01 |
| ALKINWYKK | HLA-A*03:01, HLA-A*30:01 |
| AMCTNTFVLK | HLA-A*03:01 |
| CTNTFVLKK | HLA-A*03:01, HLA-A*11:01 |
| ETQHGTILI | HLA-A*68:02 |
| ETQHGTILIK | HLA-A*68:01 |
| EVSETQHGT | HLA-A*68:02 |
| EVSETQHGTI | HLA-A*68:02 |
| GDNALKINW | HLA-B*44:02, HLA-B*44:03 |
| GEDAPCKIPF | HLA-B*40:01, HLA-B*44:02, HLA-B*44:03 |
| GESNIVIGI | HLA-B*40:01, HLA-B*44:02, HLA-B*44:03 |
| GTILIKVEY | HLA-A*01:01, HLA-A*11:01, HLA-A*26:01, HLA-A*30:02, HLA-A*32:01, HLA-B*15:01, HLA-B*57:01, HLA-B*58:01 |
| GTILIKVEYK | HLA-A*11:01 |
| HGTILIKVEY | HLA-A*26:01 |
| IGDNALKINW | HLA-B*57:01, HLA-B*58:01 |
| ITANPVVTK | HLA-A*03:01, HLA-A*11:01, HLA-A*30:01, HLA-A*68:01 |
| ITANPVVTKK | HLA-A*03:01, HLA-A*11:01, HLA-A*30:01, HLA-A*68:01 |
| KEEPVNIEA | HLA-B*40:01 |
| LITANPVVTK | HLA-A*03:01, HLA-A*11:01, HLA-A*68:01 |
| MCTNTFVLKK | HLA-A*03:01, HLA-A*11:01 |
| MSYAMCTNTF | HLA-A*23:01, HLA-A*24:02, HLA-B*57:01, HLA-B*58:01 |
| NALKINWYK | HLA-A*11:01, HLA-A*33:01, HLA-A*68:01 |
| NTFVLKKEV | HLA-A*68:02 |
| RLITANPVV | HLA-A*02:01, HLA-A*02:03, HLA-A*02:06 |
| SETQHGTIL | HLA-B*40:01, HLA-B*44:02, HLA-B*44:03 |
| SETQHGTILI | HLA-B*40:01, HLA-B*44:02 |
| SYAMCTNTF | HLA-A*23:01, HLA-A*24:02 |
| TANPVVTKK | HLA-A*03:01, HLA-A*11:01, HLA-A*30:01, HLA-A*68:01 |
| TILIKVEYK | HLA-A*03:01, HLA-A*11:01 |
| TQHGTILIK | HLA-A*03:01, HLA-A*11:01 |
| VLKKEVSET | HLA-A*02:03 |
| VSETQHGTIL | HLA-B*40:01 |

**Supplementary Table 5d: Predictions for the Serotype 4 construct sequence**

| **MHCI epitope** | **HLA allele** |
| --- | --- |
| AENTNSVTSI | HLA-B*40:01, HLA-B*44:02, HLA-B*44:03 |
| AETQHGTTV | HLA-B*40:01, HLA-B*44:02, HLA-B*44:03 |
| AETQHGTTVV | HLA-B*40:01 |
| ALTLHWFRK | HLA-A*03:01 |
| APCKVPIEI | HLA-B*07:02, HLA-B*51:01, HLA-B*53:01 |
| DSALTLHWF | HLA-A*26:01 |
| DSALTLHWFR | HLA-A*33:01, HLA-A*68:01 |
| DVNKEKVVGR | HLA-A*33:01, HLA-A*68:01 |
| EIRDVNKEK | HLA-A*68:01 |
| ELEPPFGDSY | HLA-A*01:01, HLA-A*26:01, HLA-B*44:02, HLA-B*44:03 |
| ETQHGTTVV | HLA-A*68:02 |
| ETQHGTTVVK | HLA-A*68:01 |
| GDSALTLHW | HLA-B*44:02, HLA-B*44:03 |
| GTTVVKVKY | HLA-A*01:01, HLA-A*30:02 |
| KEKVVGRII | HLA-B*44:02 |
| LEPPFGDSY | HLA-B*44:02, HLA-B*44:03 |
| NTNSVTSIEL | HLA-A*68:02 |
| RIISSTPFA | HLA-A*02:06, HLA-A*30:01 |
| SALTLHWFR | HLA-A*31:01, HLA-A*33:01, HLA-A*68:01 |
| SYTMCSGKF | HLA-A*23:01, HLA-A*24:02 |
| TPFAENTNSV | HLA-B*51:01 |
| TQHGTTVVK | HLA-A*11:01, HLA-A*30:01 |
| TSIELEPPF | HLA-B*35:01, HLA-B*53:01, HLA-B*58:01 |
| VGDSALTLHW | HLA-B*57:01, HLA-B*58:01 |
| VNKEKVVGR | HLA-A*31:01, HLA-A*33:01 |

**Supplementary Table 6a-d: Predicted T-cell epitopes (MHCII) and the corresponding HLA alleles**

We predicted the MHCII epitopes from the four serotype EDIII construct sequence using combination of NetMHCIIpan 4.0, NN-align 2.3 and SMMalign (MHCII) implemented in IEDB.

**Supplementary Table 6a: Predictions for the Serotype 1 construct sequence**

| **MHCII epitope** | **HLA allele** |
| --- | --- |
| ESYIVIGAGEKALKL | HLA-DRB5*01:01 |
| FGESYIVIGAGEKAL | HLA-DRB5*01:01 |
| GESYIVIGAGEKALK | HLA-DRB5*01:01 |
| GVSYVMCTGSFKLEK | HLA-DRB5*01:01 |
| KGVSYVMCTGSFKLE | HLA-DRB5*01:01 |
| PFGESYIVIGAGEKA | HLA-DRB5*01:01 |
| PPFGESYIVIGAGEK | HLA-DRB5*01:01 |
| SYIVIGAGEKALKLS | HLA-DRB5*01:01 |
| SYVMCTGSFKLEKEV | HLA-DRB5*01:01 |
| VSYVMCTGSFKLEKE | HLA-DRB5*01:01 |
| YIVIGAGEKALKLSW | HLA-DRB5*01:01 |
| YVMCTGSFKLEKEVA | HLA-DRB5*01:01 |
| AETQHGTVLVQVKYE | HLA-DQA1*01:02/DQB1*06:02 |
| AGEKALKLSWFKKGS | HLA-DPA1*02:01/DPB1*05:01 |
| ANPIVTDKEKPVNIE | HLA-DRB1*13:02, HLA-DRB3*01:01 |
| DKEKPVNIETEPPFG | HLA-DQA1*03:01/DQB1*03:02 |
| EKPVNIETEPPFGES | HLA-DQA1*03:01/DQB1*03:02 |
| EPPFGESYIVIGAGE | HLA-DPA1*01:03/DPB1*02:01 |
| ESYIVIGAGEKALKL | HLA-DQA1*05:01/DQB1*03:01 |
| ETEPPFGESYIVIGA | HLA-DPA1*01:03/DPB1*02:01 |
| ETQHGTVLVQVKYEG | HLA-DQA1*01:02/DQB1*06:02 |
| EVAETQHGTVLVQVK | HLA-DQA1*01:02/DQB1*06:02 |
| FGESYIVIGAGEKAL | HLA-DQA1*05:01/DQB1*03:01 |
| GAGEKALKLSWFKKG | HLA-DPA1*02:01/DPB1*05:01 |
| GEKALKLSWFKKGSS | HLA-DPA1*02:01/DPB1*05:01 |
| GESYIVIGAGEKALK | HLA-DQA1*05:01/DQB1*03:01 |
| GRLITANPIVTDKEK | HLA-DQA1*04:01/DQB1*04:02, HLA-DRB1*13:02, HLA-DRB3*02:02 |
| GVSYVMCTGSFKLEK | HLA-DRB1*07:01 |
| GVTQNGRLITANPIV | HLA-DQA1*05:01/DQB1*03:01, HLA-DRB1*13:02, HLA-DRB3*02:02, HLA-DRB4*01:01 |
| IETEPPFGESYIVIG | HLA-DPA1*01:03/DPB1*02:01 |
| IGAGEKALKLSWFKK | HLA-DPA1*02:01/DPB1*05:01 |
| IVIGAGEKALKLSWF | HLA-DQA1*05:01/DQB1*03:01 |
| IVTDKEKPVNIETEP | HLA-DQA1*03:01/DQB1*03:02 |
| KEKPVNIETEPPFGE | HLA-DQA1*03:01/DQB1*03:02 |
| KEVAETQHGTVLVQV | HLA-DQA1*01:02/DQB1*06:02 |
| KGVSYVMCTGSFKLE | HLA-DRB1*07:01 |
| KGVTQNGRLITANPI | HLA-DRB1*13:02 |
| LITANPIVTDKEKPV | HLA-DRB1*13:02 |
| NGRLITANPIVTDKE | HLA-DPA1*02:01/DPB1*14:01, HLA-DQA1*04:01/DQB1*04:02, HLA-DRB1*13:02, HLA-DRB3*02:02 |
| NIETEPPFGESYIVI | HLA-DPA1*01:03/DPB1*02:01 |
| NPIVTDKEKPVNIET | HLA-DRB1*13:02 |
| PFGESYIVIGAGEKA | HLA-DQA1*05:01/DQB1*03:01 |
| PIVTDKEKPVNIETE | HLA-DQA1*03:01/DQB1*03:02 |
| QHGTVLVQVKYEGTD | HLA-DQA1*01:02/DQB1*06:02 |
| QNGRLITANPIVTDK | HLA-DPA1*02:01/DPB1*14:01, HLA-DQA1*04:01/DQB1*04:02, HLA-DRB1*13:02, HLA-DRB3*02:02, HLA-DRB4*01:01 |
| RLITANPIVTDKEKP | HLA-DQA1*04:01/DQB1*04:02, HLA-DRB3*02:02 |
| SYIVIGAGEKALKLS | HLA-DQA1*05:01/DQB1*03:01 |
| SYVMCTGSFKLEKEV | HLA-DRB1*07:01 |
| TANPIVTDKEKPVNI | HLA-DRB1*13:02 |
| TDKEKPVNIETEPPF | HLA-DQA1*03:01/DQB1*03:02 |
| TEPPFGESYIVIGAG | HLA-DPA1*01:03/DPB1*02:01 |
| TQHGTVLVQVKYEGT | HLA-DQA1*01:02/DQB1*06:02 |
| TQNGRLITANPIVTD | HLA-DPA1*02:01/DPB1*14:01, HLA-DQA1*04:01/DQB1*04:02, HLA-DRB1*13:02, HLA-DRB3*02:02, HLA-DRB4*01:01 |
| VAETQHGTVLVQVKY | HLA-DQA1*01:02/DQB1*06:02 |
| VSYVMCTGSFKLEKE | HLA-DRB1*07:01 |
| VTDKEKPVNIETEPP | HLA-DQA1*03:01/DQB1*03:02 |
| VTQNGRLITANPIVT | HLA-DPA1*02:01/DPB1*14:01, HLA-DRB1*13:02, HLA-DRB3*02:02, HLA-DRB4*01:01 |
| YIVIGAGEKALKLSW | HLA-DQA1*05:01/DQB1*03:01 |

**Supplementary Table 6b: Predictions for the Serotype 2 construct sequence**

| **MHCII epitope** | **HLA allele** |
| --- | --- |
| DSPVNIEAEPPFGDS | HLA-DQA1*03:01, HLA-DQB1*03:02 |
| EKDSPVNIEAEPPFG | HLA-DQA1*03:01, HLA-DQB1*03:02 |
| IVTEKDSPVNIEAEP | HLA-DQA1*03:01, HLA-DQB1*03:02 |
| KDSPVNIEAEPPFGD | HLA-DQA1*03:01, HLA-DQB1*03:02 |
| PIVTEKDSPVNIEAE | HLA-DQA1*03:01, HLA-DQB1*03:02 |
| TEKDSPVNIEAEPPF | HLA-DQA1*03:01, HLA-DQB1*03:02 |
| VTEKDSPVNIEAEPP | HLA-DQA1*03:01, HLA-DQB1*03:02 |
| AEPPFGDSYIIIGVE | HLA-DQA1*04:01/DQB1*04:02 |
| AETQHGTIVVRVQYE | HLA-DQA1*01:02/DQB1*06:02 |
| CKIPFEIMDLEKRHV | HLA-DPA1*02:01/DPB1*05:01, HLA-DPA1*03:01/DPB1*04:02 |
| DLEKRHVLGRLITVN | HLA-DPA1*02:01/DPB1*14:01, HLA-DRB1*07:01 |
| DSYIIIGVEPGQLKL | HLA-DQA1*03:01/DQB1*03:02, HLA-DQA1*05:01/DQB1*02:01, HLA-DRB1*04:01, HLA-DRB1*08:02, HLA-DRB1*13:02 |
| EIAETQHGTIVVRVQ | HLA-DQA1*01:02/DQB1*06:02 |
| EIMDLEKRHVLGRLI | HLA-DRB1*07:01, HLA-DRB1*11:01 |
| EKDSPVNIEAEPPFG | HLA-DQA1*04:01/DQB1*04:02 |
| EKRHVLGRLITVNPI | HLA-DPA1*02:01/DPB1*14:01, HLA-DRB1*07:01 |
| EPGQLKLSWFKKGSS | HLA-DPA1*02:01/DPB1*05:01 |
| EPPFGDSYIIIGVEP | HLA-DQA1*04:01/DQB1*04:02 |
| ETQHGTIVVRVQYEG | HLA-DQA1*01:02/DQB1*06:02 |
| FEIMDLEKRHVLGRL | HLA-DRB1*11:01 |
| FGDSYIIIGVEPGQL | HLA-DQA1*03:01/DQB1*03:02, HLA-DQA1*04:01/DQB1*04:02, HLA-DQA1*05:01/DQB1*02:01, HLA-DRB1*13:02 |
| GDSYIIIGVEPGQLK | HLA-DQA1*03:01/DQB1*03:02, HLA-DQA1*05:01/DQB1*02:01, HLA-DRB1*04:01, HLA-DRB1*08:02, HLA-DRB1*13:02 |
| GKFKVVKEIAETQHG | HLA-DPA1*02:01/DPB1*14:01, HLA-DRB1*04:01, HLA-DRB1*08:02 |
| GMSYSMCTGKFKVVK | HLA-DRB5*01:01 |
| GRLITVNPIVTEKDS | HLA-DPA1*02:01/DPB1*14:01, HLA-DQA1*04:01/DQB1*04:02, HLA-DRB1*04:05, HLA-DRB1*08:02, HLA-DRB1*13:02, HLA-DRB3*02:02 |
| GVEPGQLKLSWFKKG | HLA-DPA1*02:01/DPB1*05:01 |
| HGTIVVRVQYEGDGS | HLA-DQA1*03:01/DQB1*03:02 |
| HVLGRLITVNPIVTE | HLA-DPA1*02:01/DPB1*14:01, HLA-DPA1*03:01/DPB1*04:02, HLA-DQA1*04:01/DQB1*04:02, HLA-DRB1*08:02, HLA-DRB1*13:02, HLA-DRB3*02:02, HLA-DRB4*01:01 |
| IAETQHGTIVVRVQY | HLA-DQA1*01:02/DQB1*06:02 |
| IMDLEKRHVLGRLIT | HLA-DRB1*07:01, HLA-DRB1*11:01 |
| IPFEIMDLEKRHVLG | HLA-DPA1*03:01/DPB1*04:02, HLA-DRB1*11:01 |
| IVTEKDSPVNIEAEP | HLA-DQA1*04:01/DQB1*04:02 |
| KEIAETQHGTIVVRV | HLA-DQA1*01:02/DQB1*06:02 |
| KGMSYSMCTGKFKVV | HLA-DRB5*01:01 |
| KIPFEIMDLEKRHVL | HLA-DPA1*02:01/DPB1*05:01, HLA-DPA1*03:01/DPB1*04:02, HLA-DRB1*11:01, HLA-DRB5*01:01 |
| KRHVLGRLITVNPIV | HLA-DPA1*02:01/DPB1*14:01, HLA-DRB1*07:01, HLA-DRB4*01:01 |
| LEKRHVLGRLITVNP | HLA-DPA1*02:01/DPB1*14:01, HLA-DRB1*07:01 |
| LGRLITVNPIVTEKD | HLA-DPA1*02:01/DPB1*14:01, HLA-DQA1*04:01/DQB1*04:02, HLA-DRB1*04:05, HLA-DRB1*08:02, HLA-DRB1*13:02, HLA-DRB3*02:02 |
| MDLEKRHVLGRLITV | HLA-DRB1*07:01 |
| MSYSMCTGKFKVVKE | HLA-DRB5*01:01 |
| NPIVTEKDSPVNIEA | HLA-DRB1*13:02 |
| PCKIPFEIMDLEKRH | HLA-DPA1*03:01/DPB1*04:02 |
| PFEIMDLEKRHVLGR | HLA-DRB1*11:01 |
| PFGDSYIIIGVEPGQ | HLA-DQA1*03:01/DQB1*03:02, HLA-DQA1*04:01/DQB1*04:02 |
| PIVTEKDSPVNIEAE | HLA-DQA1*04:01/DQB1*04:02, HLA-DRB1*13:02 |
| PPFGDSYIIIGVEPG | HLA-DQA1*03:01/DQB1*03:02, HLA-DQA1*04:01/DQB1*04:02 |
| RHVLGRLITVNPIVT | HLA-DPA1*02:01/DPB1*14:01, HLA-DPA1*03:01/DPB1*04:02, HLA-DRB1*08:02, HLA-DRB1*13:02, HLA-DRB3*02:02, HLA-DRB4*01:01 |
| RLITVNPIVTEKDSP | HLA-DQA1*04:01/DQB1*04:02, HLA-DRB3*02:02 |
| SPCKIPFEIMDLEKR | HLA-DPA1*03:01/DPB1*04:02 |
| SYIIIGVEPGQLKLS | HLA-DPA1*02:01/DPB1*14:01, HLA-DRB1*08:02, HLA-DRB1*13:02 |
| SYSMCTGKFKVVKEI | HLA-DRB5*01:01 |
| TEKDSPVNIEAEPPF | HLA-DQA1*04:01/DQB1*04:02 |
| TGKFKVVKEIAETQH | HLA-DPA1*02:01/DPB1*14:01, HLA-DRB1*04:01, HLA-DRB1*08:02 |
| TQHGTIVVRVQYEGD | HLA-DQA1*01:02/DQB1*06:02 |
| TVNPIVTEKDSPVNI | HLA-DRB1*13:02 |
| VEPGQLKLSWFKKGS | HLA-DPA1*02:01/DPB1*05:01 |
| VLGRLITVNPIVTEK | HLA-DPA1*02:01/DPB1*14:01, HLA-DQA1*04:01/DQB1*04:02, HLA-DRB1*08:02, HLA-DRB1*13:02, HLA-DRB3*02:02, HLA-DRB4*01:01, HLA-DRB1*13:02 |
| VNPIVTEKDSPVNIE | HLA-DRB1*13:02 |
| VTEKDSPVNIEAEPP | HLA-DQA1*04:01/DQB1*04:02 |
| YIIIGVEPGQLKLSW | HLA-DRB1*08:02, HLA-DRB1*13:02 |
| YSMCTGKFKVVKEIA | HLA-DRB5*01:01 |

**Supplementary Table 6c: Predictions for the Serotype 3 construct sequence**

| **MHCII epitope** | **HLA allele** |
| --- | --- |
| NGRLITANPVVTKK | HLA-DRB3*02:02 |
| GRLITANPVVTKK | HLA-DRB3*02:02 |
| GRLITANPVVTKKE | HLA-DRB3*02:02 |
| HNGRLITANPVVTKK | HLA-DRB3*02:02 |
| NGRLITANPVVTKKE | HLA-DRB3*02:02 |
| GRLITANPVVTKKEE | HLA-DRB3*02:02 |
| AHNGRLITANPVVTKK | HLA-DRB3*02:02 |
| HNGRLITANPVVTKKE | HLA-DRB3*02:02 |
| NGRLITANPVVTKKEE | HLA-DRB3*02:02 |
| HNGRLITANPVVTK | HLA-DRB3*02:02 |
| NGRLITANPVVTK | HLA-DRB3*02:02 |
| RLITANPVVTKKE | HLA-DRB3*02:02 |
| AHNGRLITANPVVTK | HLA-DRB3*02:02 |
| RLITANPVVTKK | HLA-DRB3*02:02 |
| RLITANPVVTKKEE | HLA-DRB3*02:02 |
| GRLITANPVVTKKEEP | HLA-DRB3*02:02 |
| GRLITANPVVTK | HLA-DRB3*02:02 |
| AHNGRLITANPVVTKKE | HLA-DRB3*02:02 |
| KAHNGRLITANPVVTKK | HLA-DRB3*02:02 |
| HNGRLITANPVVTKKEE | HLA-DRB3*02:02 |
| NGRLITANPVVTKKEEP | HLA-DRB3*02:02 |
| AHNGRLITANPVVTKKEE | HLA-DRB3*02:02 |
| HNGRLITANPVVTKKEEP | HLA-DRB3*02:02 |
| KAHNGRLITANPVVTKKE | HLA-DRB3*02:02 |
| GKAHNGRLITANPVVTKK | HLA-DRB3*02:02 |
| SYAMCTNTFVLK | HLA-DRB3*02:02 |
| AHNGRLITANPVV | HLA-DRB3*02:02 |
| AHNGRLITANPVVT | HLA-DPA1*02:01/DPB1*14:01, HLA-DRB3*02:02 |
| AHNGRLITANPVVTK | HLA-DPA1*02:01/DPB1*14:01 |
| AHNGRLITANPVVTKK | HLA-DPA1*02:01/DPB1*14:01 |
| AHNGRLITANPVVTKKE | HLA-DPA1*02:01/DPB1*14:01 |
| AHNGRLITANPVVTKKEE | HLA-DPA1*02:01/DPB1*14:01 |
| ALKINWYKKGSS | HLA-DRB3*02:02 |
| GKAHNGRLITANPVVT | HLA-DRB3*02:02 |
| GKAHNGRLITANPVVTK | HLA-DPA1*02:01/DPB1*14:01, HLA-DRB3*02:02 |
| GKAHNGRLITANPVVTKK | HLA-DPA1*02:01/DPB1*14:01 |
| GMSYAMCTNTFV | HLA-DRB3*02:02 |
| GMSYAMCTNTFVL | HLA-DRB3*02:02 |
| GMSYAMCTNTFVLK | HLA-DRB3*02:02 |
| GMSYAMCTNTFVLKK | HLA-DRB3*02:02 |
| GMSYAMCTNTFVLKKE | HLA-DRB3*02:02 |
| GMSYAMCTNTFVLKKEV | HLA-DRB3*02:02 |
| GMSYAMCTNTFVLKKEVS | HLA-DRB3*02:02 |
| GRLITANPVVTK | HLA-DPA1*02:01/DPB1*14:01 |
| GRLITANPVVTKK | HLA-DPA1*02:01/DPB1*14:01 |
| GRLITANPVVTKKE | HLA-DPA1*02:01/DPB1*14:01 |
| GRLITANPVVTKKEE | HLA-DPA1*02:01/DPB1*14:01 |
| GRLITANPVVTKKEEP | HLA-DPA1*02:01/DPB1*14:01 |
| GRLITANPVVTKKEEPV | HLA-DPA1*02:01/DPB1*14:01, HLA-DRB3*02:02 |
| GRLITANPVVTKKEEPVN | HLA-DRB3*02:02 |
| HNGRLITANPVV | HLA-DPA1*02:01/DPB1*14:01, HLA-DRB3*02:02 |
| HNGRLITANPVVT | HLA-DPA1*02:01/DPB1*14:01, HLA-DRB3*02:02 |
| HNGRLITANPVVTK | HLA-DPA1*02:01/DPB1*14:01 |
| HNGRLITANPVVTKK | HLA-DPA1*02:01/DPB1*14:01 |
| HNGRLITANPVVTKKE | HLA-DPA1*02:01/DPB1*14:01 |
| HNGRLITANPVVTKKEE | HLA-DPA1*02:01/DPB1*14:01 |
| HNGRLITANPVVTKKEEP | HLA-DPA1*02:01/DPB1*14:01 |
| KAHNGRLITANPVV | HLA-DRB3*02:02 |
| KAHNGRLITANPVVT | HLA-DRB3*02:02 |
| KAHNGRLITANPVVTK | HLA-DPA1*02:01/DPB1*14:01, HLA-DRB3*02:02 |
| KAHNGRLITANPVVTKK | HLA-DPA1*02:01/DPB1*14:01 |
| KAHNGRLITANPVVTKKE | HLA-DPA1*02:01/DPB1*14:01 |
| KGMSYAMCTNTFV | HLA-DRB3*02:02 |
| KGMSYAMCTNTFVL | HLA-DRB3*02:02 |
| KGMSYAMCTNTFVLK | HLA-DRB3*02:02 |
| KGMSYAMCTNTFVLKK | HLA-DRB3*02:02 |
| KGMSYAMCTNTFVLKKE | HLA-DRB3*02:02 |
| KGMSYAMCTNTFVLKKEV | HLA-DRB3*02:02 |
| LITANPVVTKKE | HLA-DRB3*02:02 |
| LITANPVVTKKEE | HLA-DRB3*02:02 |
| LITANPVVTKKEEP | HLA-DRB3*02:02 |
| LITANPVVTKKEEPV | HLA-DRB3*02:02 |
| MSYAMCTNTFVL | HLA-DRB3*02:02 |
| MSYAMCTNTFVLK | HLA-DRB3*02:02 |
| MSYAMCTNTFVLKK | HLA-DRB3*02:02 |
| MSYAMCTNTFVLKKE | HLA-DRB3*02:02 |
| MSYAMCTNTFVLKKEV | HLA-DRB3*02:02 |
| MSYAMCTNTFVLKKEVS | HLA-DRB3*02:02 |
| MSYAMCTNTFVLKKEVSE | HLA-DRB3*02:02 |
| NALKINWYKKGS | HLA-DRB3*02:02 |
| NALKINWYKKGSS | HLA-DRB3*02:02 |
| NGRLITANPVVT | HLA-DPA1*02:01/DPB1*14:01, HLA-DRB3*02:02 |
| NGRLITANPVVTK | HLA-DPA1*02:01/DPB1*14:01 |
| NGRLITANPVVTKK | HLA-DPA1*02:01/DPB1*14:01 |
| NGRLITANPVVTKKE | HLA-DPA1*02:01/DPB1*14:01 |
| NGRLITANPVVTKKEE | HLA-DPA1*02:01/DPB1*14:01 |
| NGRLITANPVVTKKEEP | HLA-DPA1*02:01/DPB1*14:01 |
| NGRLITANPVVTKKEEPV | HLA-DPA1*02:01/DPB1*14:01, HLA-DRB3*02:02 |
| QGKAHNGRLITANPVVT | HLA-DRB3*02:02 |
| QGKAHNGRLITANPVVTK | HLA-DRB3*02:02 |
| RLITANPVVTKK | HLA-DPA1*02:01/DPB1*14:01 |
| RLITANPVVTKKEEP | HLA-DRB3*02:02 |
| RLITANPVVTKKEEPV | HLA-DRB3*02:02 |
| RLITANPVVTKKEEPVN | HLA-DRB3*02:02 |
| RLITANPVVTKKEEPVNI | HLA-DRB3*02:02 |
| SYAMCTNTFVLK | HLA-DPA1*01:03/DPB1*04:01 |
| SYAMCTNTFVLKK | HLA-DPA1*01:03/DPB1*04:01, HLA-DRB3*02:02 |
| SYAMCTNTFVLKKE | HLA-DRB3*02:02 |
| SYAMCTNTFVLKKEV | HLA-DRB3*02:02 |
| SYAMCTNTFVLKKEVS | HLA-DRB3*02:02 |
| SYAMCTNTFVLKKEVSE | HLA-DRB3*02:02 |
| YAMCTNTFVLKK | HLA-DPA1*01:03/DPB1*04:01, HLA-DRB3*02:02 |
| YAMCTNTFVLKKE | HLA-DRB3*02:02 |
| YAMCTNTFVLKKEV | HLA-DRB3*02:02, |

**Supplementary Table 6d: Predictions for the Serotype 4 construct sequence**

| **MHCII epitope** | **HLA allele** |
| --- | --- |
| VGRIISSTPFAENT | HLA-DPA1*02:01, HLA-DPB1*14:01 |
| SSTPFAENTNSVTSI | HLA-DRB3*02:02 |
| STPFAENTNSVTSIE | HLA-DRB3*02:02 |
| ISSTPFAENTNSVTSI | HLA-DRB3*02:02 |
| SSTPFAENTNSVTSIE | HLA-DRB3*02:02 |
| STPFAENTNSVTSIEL | HLA-DRB3*02:02 |
| VVGRIISSTPFAENT | HLA-DPA1*02:01, HLA-DPB1*14:01 |
| STPFAENTNSVTSI | HLA-DRB3*02:02 |
| VVGRIISSTPFAEN | HLA-DPA1*02:01, HLA-DPB1*14:01 |
| GRIISSTPFAENT | HLA-DPA1*02:01, HLA-DPB1*14:01 |
| VGRIISSTPFAEN | HLA-DPA1*02:01, HLA-DPB1*14:01 |
| IISSTPFAENTNSVTSI | HLA-DRB3*02:02 |
| ISSTPFAENTNSVTSIE | HLA-DRB3*02:02 |
| SSTPFAENTNSVTSIEL | HLA-DRB3*02:02 |
| STPFAENTNSVTSIELE | HLA-DRB3*02:02 |
| GRIISSTPFAEN | HLA-DPA1*02:01, HLA-DPB1*14:01 |
| IISSTPFAENTNSVTSIE | HLA-DRB3*02:02 |
| ISSTPFAENTNSVTSIEL | HLA-DRB3*02:02 |
| RIISSTPFAENTNSVTSI | HLA-DRB3*02:02 |
| SSTPFAENTNSVTSIELE | HLA-DRB3*02:02 |
| RIISSTPFAENT | HLA-DPA1*02:01, HLA-DPB1*14:01 |
| VGRIISSTPFAE | HLA-DPA1*02:01, HLA-DPB1*14:01 |
| DVNKEKVVGRIISSTPFA | HLA-DPA1*02:01/DPB1*14:01 |
| EKVVGRIISSTPFA | HLA-DPA1*02:01/DPB1*14:01 |
| EKVVGRIISSTPFAE | HLA-DPA1*02:01/DPB1*14:01 |
| EKVVGRIISSTPFAEN | HLA-DPA1*01:03/DPB1*04:01, HLA-DPA1*02:01/DPB1*14:01 |
| EKVVGRIISSTPFAENT | HLA-DPA1*01:03/DPB1*04:01, HLA-DPA1*02:01/DPB1*14:01 |
| EKVVGRIISSTPFAENTN | HLA-DPA1*01:03/DPB1*04:01, HLA-DPA1*02:01/DPB1*14:01 |
| GRIISSTPFAEN | HLA-DPA1*01:03/DPB1*04:01, HLA-DRB3*02:02 |
| GRIISSTPFAENT | HLA-DPA1*01:03/DPB1*04:01, HLA-DRB3*02:02 |
| GRIISSTPFAENTN | HLA-DPA1*01:03/DPB1*04:01, HLA-DPA1*02:01/DPB1*14:01, HLA-DRB3*02:02 |
| GRIISSTPFAENTNS | HLA-DPA1*01:03/DPB1*04:01, HLA-DPA1*02:01/DPB1*14:01 |
| GRIISSTPFAENTNSV | HLA-DPA1*01:03/DPB1*04:01, HLA-DPA1*02:01/DPB1*14:01 |
| GRIISSTPFAENTNSVT | HLA-DPA1*01:03/DPB1*04:01, HLA-DPA1*02:01/DPB1*14:01, HLA-DRB3*02:02 |
| GRIISSTPFAENTNSVTS | HLA-DPA1*01:03/DPB1*04:01, HLA-DRB3*02:02 |
| IISSTPFAENTNSVT | HLA-DRB3*02:02 |
| IISSTPFAENTNSVTS | HLA-DRB3*02:02 |
| ISSTPFAENTNSVT | HLA-DRB3*02:02 |
| ISSTPFAENTNSVTS | HLA-DRB3*02:02 |
| KEKVVGRIISST | HLA-DPA1*02:01/DPB1*14:01 |
| KEKVVGRIISSTP | HLA-DPA1*02:01/DPB1*14:01 |
| KEKVVGRIISSTPF | HLA-DPA1*02:01/DPB1*14:01 |
| KEKVVGRIISSTPFA | HLA-DPA1*02:01/DPB1*14:01 |
| KEKVVGRIISSTPFAE | HLA-DPA1*02:01/DPB1*14:01 |
| KEKVVGRIISSTPFAEN | HLA-DPA1*01:03/DPB1*04:01, HLA-DPA1*02:01/DPB1*14:01 |
| KEKVVGRIISSTPFAENT | HLA-DPA1*01:03/DPB1*04:01, HLA-DPA1*02:01/DPB1*14:01 |
| KVVGRIISSTPFA | HLA-DPA1*02:01/DPB1*14:01 |
| KVVGRIISSTPFAE | HLA-DPA1*02:01/DPB1*14:01, HLA-DRB3*02:02 |
| KVVGRIISSTPFAEN | HLA-DPA1*01:03/DPB1*04:01, HLA-DPA1*02:01/DPB1*14:01, HLA-DRB3*02:02 |
| KVVGRIISSTPFAENT | HLA-DPA1*01:03/DPB1*04:01, HLA-DPA1*02:01/DPB1*14:01, HLA-DRB3*02:02 |
| KVVGRIISSTPFAENTN | HLA-DPA1*01:03/DPB1*04:01, HLA-DPA1*02:01/DPB1*14:01 |
| KVVGRIISSTPFAENTNS | HLA-DPA1*01:03/DPB1*04:01, HLA-DPA1*02:01/DPB1*14:01 |
| NKEKVVGRIISS | HLA-DPA1*02:01/DPB1*14:01 |
| NKEKVVGRIISST | HLA-DPA1*02:01/DPB1*14:01 |
| NKEKVVGRIISSTPF | HLA-DPA1*02:01/DPB1*14:01 |
| NKEKVVGRIISSTPFA | HLA-DPA1*02:01/DPB1*14:01 |
| NKEKVVGRIISSTPFAE | HLA-DPA1*02:01/DPB1*14:01 |
| NKEKVVGRIISSTPFAEN | HLA-DPA1*01:03/DPB1*04:01, HLA-DPA1*02:01/DPB1*14:01 |
| PFAENTNSVTSI | HLA-DRB3*02:02 |
| PFAENTNSVTSIE | HLA-DRB3*02:02 |
| PFAENTNSVTSIEL | HLA-DRB3*02:02 |
| PFAENTNSVTSIELE | HLA-DRB3*02:02 |
| PFAENTNSVTSIELEP | HLA-DRB3*02:02 |
| PFAENTNSVTSIELEPP | HLA-DRB3*02:02 |
| PFAENTNSVTSIELEPPF | HLA-DRB3*02:02 |
| RIISSTPFAENT | HLA-DPA1*01:03/DPB1*04:01, HLA-DRB3*02:02 |
| RIISSTPFAENTN | HLA-DPA1*01:03/DPB1*04:01, HLA-DPA1*02:01/DPB1*14:01 |
| RIISSTPFAENTNS | HLA-DPA1*01:03/DPB1*04:01, HLA-DPA1*02:01/DPB1*14:01 |
| RIISSTPFAENTNSV | HLA-DPA1*01:03/DPB1*04:01, HLA-DPA1*02:01/DPB1*14:01 |
| RIISSTPFAENTNSVT | HLA-DRB3*02:02 |
| RIISSTPFAENTNSVTS | HLA-DRB3*02:02 |
| SSTPFAENTNSVT | HLA-DRB3*02:02 |
| SSTPFAENTNSVTS | HLA-DRB3*02:02 |
| STPFAENTNSVT | HLA-DRB3*02:02 |
| STPFAENTNSVTS | HLA-DRB3*02:02 |
| STPFAENTNSVTSIELEP | HLA-DRB3*02:02 |
| TPFAENTNSVTS | HLA-DRB3*02:02 |
| TPFAENTNSVTSI | HLA-DRB3*02:02 |
| TPFAENTNSVTSIE | HLA-DRB3*02:02 |
| TPFAENTNSVTSIEL | HLA-DRB3*02:02 |
| TPFAENTNSVTSIELE | HLA-DRB3*02:02 |
| TPFAENTNSVTSIELEP | HLA-DRB3*02:02 |
| TPFAENTNSVTSIELEPP | HLA-DRB3*02:02 |
| VGRIISSTPFAE | HLA-DPA1*01:03/DPB1*04:01, HLA-DRB3*02:02 |
| VGRIISSTPFAEN | HLA-DPA1*01:03/DPB1*04:01, HLA-DRB3*02:02 |
| VGRIISSTPFAENT | HLA-DPA1*01:03/DPB1*04:01, HLA-DRB3*02:02 |
| VGRIISSTPFAENTN | HLA-DPA1*01:03/DPB1*04:01, HLA-DPA1*02:01/DPB1*14:01, HLA-DRB3*02:02 |
| VGRIISSTPFAENTNS | HLA-DPA1*01:03/DPB1*04:01, HLA-DPA1*02:01/DPB1*14:01 |
| VGRIISSTPFAENTNSV | HLA-DPA1*01:03/DPB1*04:01, HLA-DPA1*02:01/DPB1*14:01 |
| VGRIISSTPFAENTNSVT | HLA-DPA1*01:03/DPB1*04:01, HLA-DPA1*02:01/DPB1*14:01, HLA-DRB3*02:02 |
| VNKEKVVGRIISST | HLA-DPA1*02:01/DPB1*14:01 |
| VNKEKVVGRIISSTPF | HLA-DPA1*02:01/DPB1*14:01 |
| VNKEKVVGRIISSTPFA | HLA-DPA1*02:01/DPB1*14:01 |
| VNKEKVVGRIISSTPFAE | HLA-DPA1*02:01/DPB1*14:01 |
| VVGRIISSTPFA | HLA-DPA1*02:01/DPB1*14:01, HLA-DRB3*02:02 |
| VVGRIISSTPFAE | HLA-DPA1*01:03/DPB1*04:01, HLA-DPA1*02:01/DPB1*14:01, HLA-DRB3*02:02 |
| VVGRIISSTPFAEN | HLA-DPA1*01:03/DPB1*04:01, HLA-DRB3*02:02 |
| VVGRIISSTPFAENT | HLA-DPA1*01:03/DPB1*04:01, HLA-DRB3*02:02 |
| VVGRIISSTPFAENTN | HLA-DPA1*01:03/DPB1*04:01, HLA-DPA1*02:01/DPB1*14:01 |
| VVGRIISSTPFAENTNS | HLA-DPA1*01:03/DPB1*04:01, HLA-DPA1*02:01/DPB1*14:01 |
| VVGRIISSTPFAENTNSV | HLA-DPA1*01:03/DPB1*04:01, HLA-DPA1*02:01/DPB1*14:01 |

**Supplementary Table 7: World population coverage of the T-cell epitopes**

T-cell epitopes were predicted from the dengue constructs for the four serotypes and the HLA allele coverage was used to predict the world population coverage. We have presented the projected population coverage, average number of epitope hits / HLA combinations recognized by the population, and minimum number of epitope hits / HLA combinations recognized by 90% of the population (PC90).

| **Dengue serotype** | **MHC class** | **World population coverage** | **Average hit** | **PC90** |
| --- | --- | --- | --- | --- |
| 1 | I | 97.34 | 4.27 | 1.48 |
| 1 | II | 99.43 | 21.52 | 12.07 |
| 2 | I | 98.37 | 6.15 | 2.32 |
| 2 | II | 97.94 | 24.89 | 7.66 |
| 3 | I | 96.55 | 6.82 | 1.43 |
| 3 | II | 90.16 | 13.63 | 3.01 |
| 4 | I | 90.28 | 3.22 | 1.01 |
| 4 | II | 90.16 | 52.49 | 34.07 |
